# First report on the Latvian SARS-CoV-2 isolate genetic diversity

**DOI:** 10.1101/2020.09.08.20190504

**Authors:** Nikita Zrelovs, Monta Ustinova, Ivars Silamiķelis, Līga Birzniece, Kaspars Megnis, Vita Rovīte, Lauma Freimane, Laila Silamiķele, Laura Ansone, Jānis Pjalkovskis, Dāvids Fridmanis, Baiba Vilne, Marta Priedīte, Anastasija Caica, Mikus Gavars, Dmitrijs Perminovs, Jeļena Storoženko, Oksana Savicka, Elīna Dimiņa, Uga Dumpis, Jānis Kloviņš

## Abstract

Remaining a major healthcare concern with nearly 29 million confirmed cases worldwide at the time of writing, novel severe acute respiratory syndrome coronavirus - 2 (SARS-CoV-2) has caused more than 920 thousand deaths since its outbreak in China, December 2019. First case of a person testing positive for SARS-CoV-2 infection within the territory of the Republic of Latvia was registered on 2^nd^ of March 2020, nine days prior to the pandemic declaration by WHO. Since then, more than 277 000 tests were carried out confirming a total of 1464 cases of COVID-19 in the country as of 12^th^ of September 2020. Rapidly reacting to the spread of the infection, an ongoing sequencing campaign was started mid-March in collaboration with the local testing laboratories, with an ultimate goal in sequencing as much local viral isolates as possible, resulting in first full-length SARS-CoV-2 isolate genome sequences from the Baltics region being made publicly available in early April. With 133 viral isolates representing ∼9.1% of the total COVID-19 cases in the country being completely sequenced as of today, here we provide a first report on the genetic diversity of Latvian SARS-CoV-2 isolates.

## Introduction

Current novel coronavirus disease (COVID-19) pandemic caused by severe acute respiratory syndrome coronavirus 2 (SARS-CoV-2), which was formerly known as 2019 novel coronavirus (2019-nCoV), and is often referred to as Human coronavirus 2019 (hCoV-19), responsible for a sudden rise in pneumonia cases in Wuhan, China, late December 2019, was preventively deemed a Public Health Emergency of International Concern by WHO as early as 30th January, 2020 with only as few as 7836 cases confirmed worldwide back then. With rapidly growing number of confirmed positive cases throughout the world, SARS-CoV-2 quickly became arguably the most sequenced virus in history with more than 100 thousands (2020-09-14) viral isolate near complete genome sequences of high quality available publicly at the time of writing at GISAID repository thanks to the unprecedented rate of collaborations between researchers and unpublished data sharing with the goal of effectively tackling the novel disease [1,2].

### Genome of SARS-CoV-2

First reported genomic sequence of SARS-CoV-2 was deduced from a metagenomic RNA of bronchoalveolar lavage fluid specimen sampled from a patient who worked at Wuhan seafood market, where the epidemiological onset of human-to-human transmission of a novel zoonotic coronavirus is thought to have taken place [3], although evidence of an earlier contraction of the disease that was not associated with the seafood market has been documented, leading to the conclusion that the primary spill-over event has taken place elsewhere[4,5]. The sequence of a 29903 base long non-segmented positive-sense single-stranded RNA molecule representing complete genome of the aforementioned isolate Wuhan-Hu-1 was deposited in GenBank [6] on 5^th^ of January, 2020 and is now known as a SARS-CoV-2 reference sequence available under accession numbers NC_045512.2 or MN908947.3.

While viral family *Coronaviridae*, that comprises α/β/Δ/γ coronavirus genera, representatives are somewhat unique in comparison with most other RNA viruses in regards to their large genome size of ∼30 kb, genomic organization of individual species does not differ much among other lower taxa within the family, while boasting variable number of open reading frames. The genome of SARS-CoV-2 begins with a 265 base long 5’ UTR region starting with a leader sequence followed by a 21 290 base long ORF1ab, comprising about 70% of the genome length, that translates into two polyproteins via −1 ribosomal frameshift and encodes 16 non-structural proteins (nsp1-nsp16). The remaining part of the genome comprises ORFs coding for structural and accessory proteins of unknown function, sequentially: Spike glycoprotein (S), ORF3a, envelope protein (E), membrane glycoprotein (M), ORF6, ORF7a, ORF7b, ORF8, nucleocapsid phosphoprotein (N), ORF10, followed by 3’ UTR ending in poly(A) tail. However, no evidence that would support the expression of SARS-CoV-2 ORF10 encoded protein of unknown function is yet found in the literature [7].

### Possible origins of SARS-CoV-2

SARS-CoV-2 is the seventh zoonotic human coronavirus known up to date, and, along with SARS-CoV and MERS-CoV, is considered to be highly pathogenic and more severe compared to other, milder symptoms causing, community-acquired human coronaviruses (HCoV-229E, HCoV-OC43, HCoV-HKU1 and HCoV-NL63) [8].

Studies on the origin of novel coronavirus have revealed that complete genomic sequence of SARS-CoV-2 suggests a more close, although not a direct parental, ancestral relationship with bat (∼96% overall nucleotide homology with RaTG13[9]) and pangolin coronaviruses (up to ∼92% homology, with S protein ACE2 receptor binding domain amino acid sequence being 97.4% identical to SARS-CoV-2 [10]), than to those of humans(∼79% and ∼50% identity to SARS-CoV and MERS-CoV, respectively [11], and, while bats are already a long-time acknowledged reservoir of SARS-CoV-like β-coronaviruses [12,13], the assumption that pangolins can serve as a natural host for CoVs has been made only recently before the emergence of SARS-CoV-2 [14,15]. Although the current risk of animal-human transmission of COVID-19 is considered low, a number of felines[16], canines[17] and minks[18] worldwide have been reported to be infected with SARS-CoV-2.

### SARS-CoV-2 isolate classification

With a steadily growing number of complete hCoV-19 genome sequences, early efforts to classify novel isolates based on their genetic make-up have resulted in numerous proposals of different SARS-CoV-2 isolate classification systems [19][20] [21], some of which (e.g. PANGOLIN lineages) are complementary. However, with more than 100 000 of hCoV-19 genome sequences being available publicly as of now, ongoing efforts to aid in the classification of newly sequenced viral isolates have resulted in the general acceptance of GISAID’s team developed SARS-CoV-2 major clade and lineage nomenclature system based on the specific combinations of 9 hCoV-19 genetic markers [1]. In accordance with this system, hCoV-19 isolates can be classified in at least six distinct major clades, namely: S, L (containing reference sequence Wuhan-Hu-1), V, G, GH, GR and O (other) isolate clades (Table 1).

**Table 1.**
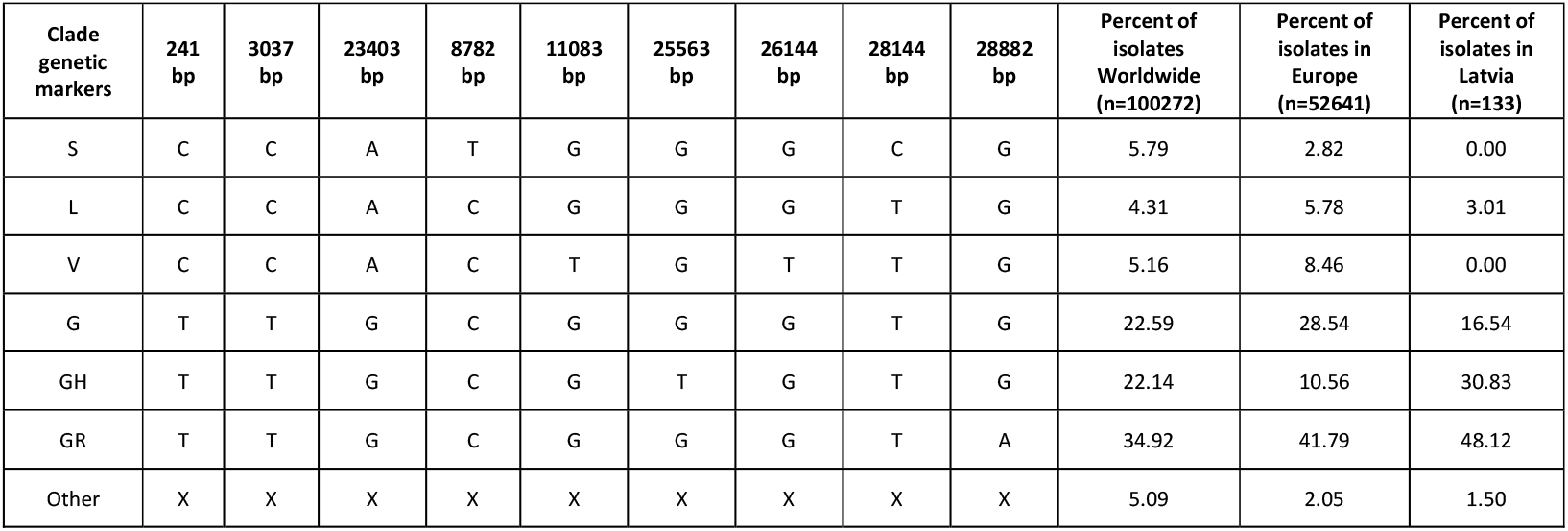
Major hCoV-19 clades defining genetic markers and their occurrence in Latvia, Europe and Worldwide (as of 2020-09-14). X denotes any nucleotide.

| Clade genetic markers | 241 bp | 3037 bp | 23403 bp | 8782 bp | 11083 bp | 25563 bp | 26144 bp | 28144 bp | 28882 bp | Percent of isolates Worldwide (n=100272) | Percent of isolates in Europe (n=52641) | Percent of isolates in Latvia (n=133) |
| --- | --- | --- | --- | --- | --- | --- | --- | --- | --- | --- | --- | --- |
| S | C | C | A | T | G | G | G | C | G | 5.79 | 2.82 | 0.00 |
| L | C | C | A | C | G | G | G | T | G | 4.31 | 5.78 | 3.01 |
| V | C | C | A | C | T | G | T | T | G | 5.16 | 8.46 | 0.00 |
| G | T | T | G | C | G | G | G | T | G | 22.59 | 28.54 | 16.54 |
| GH | T | T | G | C | G | T | G | T | G | 22.14 | 10.56 | 30.83 |
| GR | T | T | G | C | G | G | G | T | A | 34.92 | 41.79 | 48.12 |
| Other | X | X | X | X | X | X | X | X | X | 5.09 | 2.05 | 1.50 |

Mid-September, 2020, the most represented clades Worldwide are GR, G and GH, roughly corresponding to 34.92%, 22.59% and 22.14% of total hCoV-19 isolates, respectively. All three of these clades are characterized by C241T base substitution in 5’ UTR region, C3037T silent mutation in ORF1a and missense A23403G mutation that causes Aspartic acid at position 614 of spike glycoprotein (S) to change to Glycine (S-D614G), that is associated with higher viral loads and, in turn, is hypothesized to increase the infectivity of these genotypes, with isolates bearing this mutation quickly becoming dominant ones in various regions throughout the world [22–24]. More recent clades GR and GH are further distinguished from the ancestral G genotype by G25563T mutation resulting in position 57 of ORF3a protein to change from glutamine to histidine for clade GH, and G28882A that changes glycine at nucleocapsid phosphoprotein (N) aa position 204 to arginine for clade GR. While the exact effect of GH clade-defining G25563T change in apoptosis-inducing transmembrane ORF3a protein (Q57R) remains unknown, it does not seem to affect any of the conserved functional domains distinguishable within the protein [25,26]. Whereas, G28882A mutation associated with GR genotype is almost always a trinucleotide mutation of neighboring loci resulting in GGG to AAC change at positions 28881, 28882 and 28883, respectively. This trinucleotide mutation results in two (R203K and G204R) consecutive amino acid changes in N protein, which, in turn, might have potential implications on nucleocapsid phosphoprotein structure and/or function via reduction of conformational entropy and changes in inter-residue interactions in the proximity of the mutated amino acid positions (elaborated on in [27]). The currently estimated mutation rate of SARS-CoV-2 is around 9.86 × 10^−4^ to 1.85 × 10^−4^ substitutions per position per year [28], and, based on the isolates sequenced worldwide up to date, there is evidence that mutations in nearly every position in the genome of SARS-CoV-2 have already been documented [29].

In this study, we are reporting the first results of an ongoing massive sequencing campaign that allows us to elaborate on the genetic diversity of SARS-CoV-2 isolates from Latvian patients.

## Materials and Methods

### Sample management and detection of SARS-CoV-2

For viral genome analysis either oropharyngeal or nasopharyngeal swabs obtained from COVID-19 patients or already extracted RNA samples were provided to Latvian Biomedical Research and Study Centre by the three accredited diagnostic laboratories (E. Gulbis Laboratory, Central Laboratory and Latvian Centre of Infectious Diseases) covering diagnostics of all officially reported cases of SARS-CoV-2. RNA extraction from oropharyngeal and nasopharyngeal swabs and the following SARS-CoV-2 detection was performed by multiple different methods according to standard procedures of each laboratory. These included manual Trizol-based RNA extraction (TRI reagent, Sigma) and automated purification methods with STARMag 96 × 4 Universal Cartridge Kit (Seegene Inc.), NucliSENS easyMAG (bioMérieux), QIAamp 96 Virus QIAcube HT Kit (QIAGEN). The presence of SARS-CoV-2 in the purified RNA samples was estimated by, either commercial or in-house RT-QPCR (Allplex™ 2019-nCoV Assay, Seegene Inc) methods. Samples showing amplification (ct < 40) of at least one viral gene (RdRp, E, N) were considered as positive and directed to next-generation sequencing.

### Next-generation sequencing

Metatranscriptome sequencing was the first-choice method for the SARS-CoV-2 genome analysis. Nevertheless, since the majority of samples showed an insufficient number of sequencing reads mapping to the SARS-CoV-2 genome and could not be reliably analyzed, targeted sequencing approaches were considered. A methodological strategy plan was developed in order to apply the most effective next-generation sequencing method for each sample according to the quantity of SARS-CoV-2 (Supplementary figure 1). At first, RT-QPCR was repeated for each sample in order to evaluate the quantity of viral RNA with a common approach for all samples. Three SARS-CoV-2 genome-specific primer pairs and probes targeting different regions of the nucleocapsid protein (N) gene implemented in the 2019-nCoV RUO Kit (IDT), and SOLIScript® 1-step CoV Kit (Solis Biodyne) were used for the amplification. Probes N1 and N2 specifically detected SARS-CoV-2, while the N3 probe universally detected all currently recognized clade 2 and 3 viruses within the subgenus Sarbecovirus [30]. To evaluate the RNA extraction and PCR efficiency, simultaneous amplification of the human RNase P gene was performed and a control sample containing a plasmid with the SARS-CoV-2 nucleoplasmid protein gene (2019-nCoV_N_Positive Control, IDT) was added to each reaction set. The potential contamination was evaluated by a negative control (nuclease-free water instead of RNA) added to each sample set. RT-QPCR was conducted on the ViiA 7 Real-Time PCR System (Thermo Fisher Scientific), and only the samples showing amplification (ct < 40) of all three SARS-CoV-2 nucleoplasmid protein genes were further directed to metatranscriptome sequencing. Samples exhibiting poor amplification of viral genes (ct > 40 for at least one target region) were considered for one of targeted sequencing approaches: hybridization capture or amplification of SARS-CoV-2.

### Metatranscriptome sequencing

In order to eliminate contaminating DNA, DNase I treatment (NEB) of RNA samples was performed, followed by rRNA depletion with MGIEasy rRNA Depletion Kit (MGI Tech Co. Ltd). Complementary DNA libraries were prepared using MGIEasy RNA Library Prep Set (MGI Tech Co. Ltd). Quantity and quality of both RNA and cDNA were evaluated using the Qubit 2.0 fluorometer and Agilent 2100 Bioanalyzer system, respectively. The presence of the SARS-CoV-2 genome was repeatedly tested in each cDNA library by Q-PCR before sequencing, using the same primers and probes (2019-nCoV_N_Positive Control, IDT) together with TaqMan™ Gene Expression Master Mix (Thermo Fisher Scientific). After multiple experimental tests, a ct value threshold of 25 was chosen for N1 and N3 probes for cDNA libraries to be forwarded to metatranscriptome sequencing (N2 probe appeared to be unstable and therefore uninformative). Metatranscriptome cDNA libraries were sequenced on the DNBSEQ-G400RS sequencing platform with DNBSEQ-G400RS High-throughput Sequencing Set (PE150) (MGI Tech Co. Ltd), obtaining at least 100 million 150-bp-paired-end sequencing reads per each sample. Those libraries that failed to pass the ct threshold (ct > 25 for N1 and N3) were directed to a targeted approach.

### SARS-CoV-2 hybridization capture

One of the targeted sequencing strategies was based on the enrichment of the SARS-CoV-2 genome by hybridization probes. For cDNA library preparation TruSeq Stranded Total RNA Library Prep Gold kit and TruSeq RNA UD Indexes (Illumina) were used. The indexed cDNA libraries were enriched for the SARS-CoV-2 genome using compatible hybridization probes implemented in the myBaits Expert SARS-CoV-2 kit (Arbor Biosciences) according to manufacturers instructions. The enriched libraries were sequenced on Illumina MiSeq system with MiSeq Reagent Kit v3 (150-cycle), obtaining at least 1 million of around 75-bp-paired-end reads per sample.

### Amplification of SARS-CoV-2 genome

The second targeted approach involved multiplexed primers for amplification of the whole SARS-CoV-2 genome. QIAseq SARS-CoV-2 Primer Panel (QIAGEN) [31] was used together with QIAseq FX DNA Library Kit (QIAGEN) for cDNA library preparation. Next-generation sequencing was performed on Illumina MiSeq system with MiSeq Reagent Kit v2 (300-cycles), obtaining at least 1 million of around 150bp paired-end reads per sample.

### Sequencing data quality control, variant calling and data sharing

Adapter clipping was performed with cutadapt 1.16 [32]. Subsequent read trimming was performed with fastp 0.20.0 [33] using 5 base sliding window trimming from both ends with quality threshold 20. Reads with length less than 75 bp or an average quality of less than 20 were removed. Quality controlled reads were then aligned against SARS-CoV-2 isolate Wuhan-Hu-1 reference genome (Accession number: NC_045512.2) with bowtie2 2.3.5.1 [34]. Variant calling and consensus sequence construction were implemented using bcftools 1.10.2 [35]. Average coverage for each of the genomes was calculated using samtools and in-house awk [36] scripts. Less than 1% of the missing bases were allowed for a genome to be considered successfully sequenced and missing bases were treated as reference bases from the Wuhan-Hu-1 genome. Consensus sequences of the successfully sequenced isolates were then proceeded to the manual variant quality inspection by sequence alignment map visualization in IGV [37], sequences that have passed the manual variant quality check were immediately publicly shared by deposition to GISAID database [1]. Variant annotations were performed using coronapp SARS-CoV-2 genome autoannotation web server by comparisons to reference sequence [38] and the results were summarized with the help of custom R scripts, ggplot2 R library was used for plot visualizations [39][40].

### Phylogenetic reconstructions

Sequences of the Latvian SARS-CoV-2 isolates and Wuhan-Hu-1 reference sequence were aligned using Clustal-Omega v 1.2.4. [41]. Maximum likelihood phylogeny was performed using IQTREE v 2.0.6 [42] with GTR+F+I as best fit model determined by ModelFinder [43] according to Bayesian Information Criterion (ultrafast bootstrap with 1000 replicates [44]) and assessment of temporal signal associated with the data was performed by importing resulting ML tree into TempEst v1.5.3 [45], parsing sampling dates of isolates and visualizing the root-to-tip divergence.

Bayesian phylogenetic trees were estimated using BEAST v1.10.4 [46], employing GTR nucleotide substitution model with empirical base frequencies and invariant site proportion assuming strict molecular clock. Coalescent exponential growth prior (growth rate prior: Laplace with scale 100; population size prior: Lognormal with mu 1 and sigma 2) with growth rate parametrization [47,48] was selected and Markov chain Monte Carlo (MCMC) was run for 50 million states sampling log parameters and trees every 5 000 states. Tracer v 1.7.1 [49] was used for MCMC trace (log file) inspection to evaluate sufficiency of sampling (all parameters had an ESS of more than 400) and infer substitution rate along with the date of the most recent common ancestor estimate. To summarize Bayesian phylogenetic inference, maximum clade credibility time-scaled tree was generated in TreeAnnotator v 1.10.4 (distributed with BEAST package) using 10% of the states (5 million) as the burn-in and visualized using FigTree v 1.4.4 [50].

## Results and discussion

With 1464 cumulative positive cases as of 12^th^ of September 2020 (1248 people recovered, 181 active cases of the disease and 35 COVID-19 associated deaths), 133 hCoV-19 isolates representing ∼9.1% of the total local COVID-19 cases have been completely sequenced as of today, making Latvia one of the leading countries not only in regards to the containment of the spread of COVID-19 disease, but in the number of sequenced SARS-CoV-2 isolates to the cumulative number of positive COVID-19 cases ratio as well (Figure 1).

**Figure 1.**
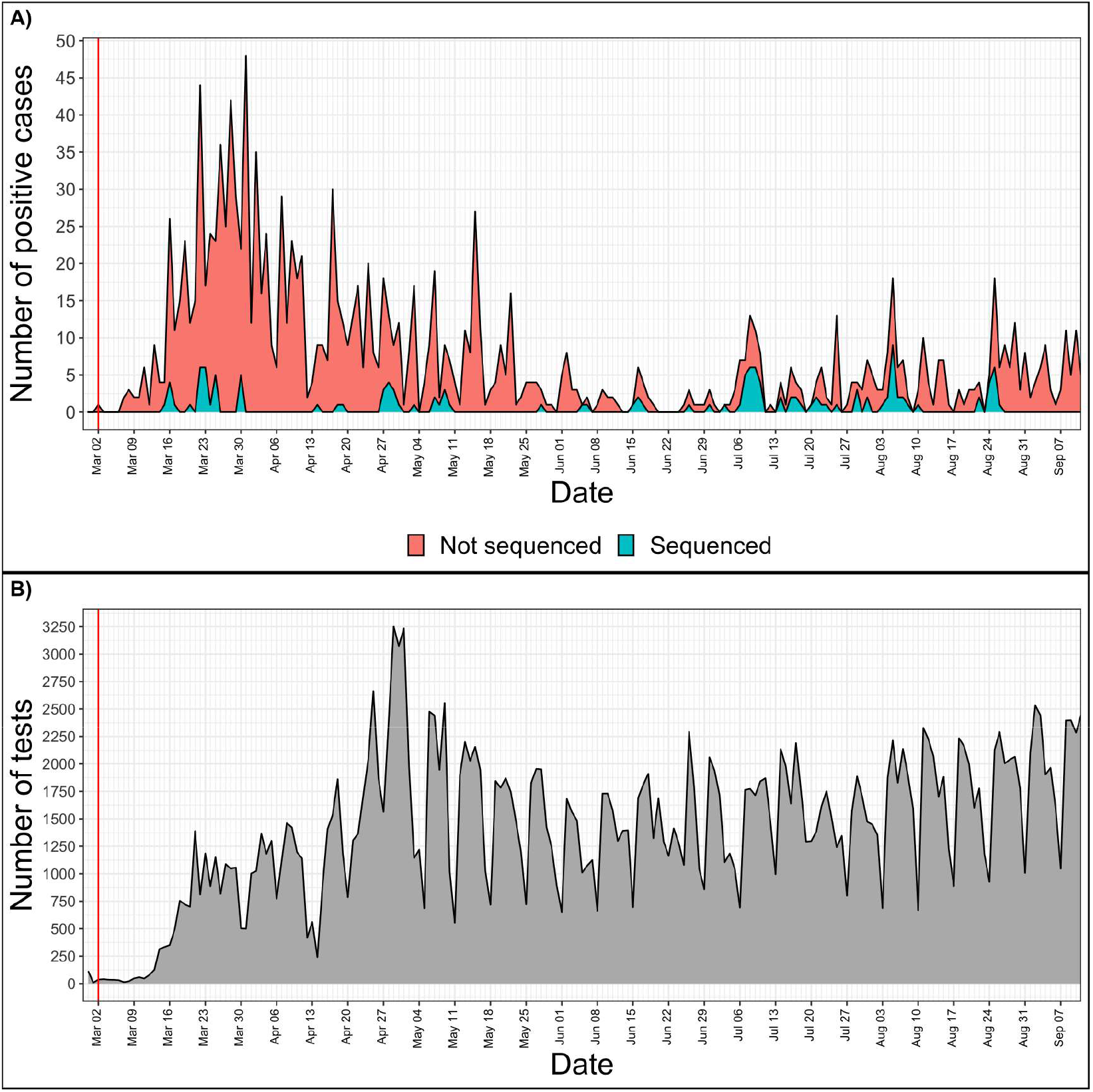
Daily numbers of positive COVID-19 cases (A) and tests performed (B) in Latvia. X axis is the same for both tiles and represents daily time series from 28^th^ of February, 2020 to 11^th^ of September, 2020. The red vertical line indicates the date of the first COVID-19 case registered in Latvia. A) Y value represents the total number of positive cases registered on a given day. Blue area shows the number of successfully sequenced isolates, while the red area represents the positive cases not sequenced during this study. B) Y value represents the number of tests carried out on a given date in Latvia.

Reacting to the emergence of the SARS-CoV-2 in Latvia, a high-throughput framework for SARS-CoV-2 isolate sequencing and data analysis with capabilities of near real-time tracking of the epidemiological situation in Latvia was built to aid the governmental decision-making and study the molecular epidemiology of hCoV19.

One of the challenges to obtain good quality sequences for maximal number of samples is the variable quality of input material that can be caused by highly variable viral loads, different collection, storage and RNA isolation methods. Although for the current study we did not have the information on the severity of COVID-19 symptoms for particular cases, it should be noted that the absolute majority of cases in Latvia are with low symptom severity expected to have lower concentration of virus in diagnostic samples. We therefore developed an approach to verify sample quality and select appropriate sequencing method to recover maximal available information from existing samples ensuring cost efficacy of the process (Figure S1). According to this strategy developed during the implementation of the study, complete SARS-CoV-2 genome sequence was successfully obtained by metatranscriptome approach for 37 viral isolates, 80 samples were analyzed by amplification of SARS-CoV-2 genome with multiplexed primers, and for 16 isolates enrichment of SARS-CoV-2 genome was performed by hybridization capture method prior the sequencing.

As of now it could be cautiously speculated that the obtained results on the SARS-CoV-2 genotype distributions might be somewhat representative of a whole Baltics region, taking the geographical proximity, travel habits and mild governmental travel regulations between the Baltic states during the most of the pandemic into the account. However, the extent of similarity between the isolates circulating in different Baltic states currently cannot be reliably established due to SARS-CoV-2 isolate undersequencing in neigboring Estonia and Lithuania, and the founder effect of multiple independent (re-)introductions of different SARS-CoV-2 genotypes, as well as containment effectivity of respective COVID19 cases, in each of the countries should not be overlooked.

### Distribution of sequenced virus isolates by SARS-CoV-2 clades

Major isolate clade distributions across distinct geographical regions show clear spatial differences of the epidemic (Figure 2) and a trend of “older” isolate clades L and S losing their initial prevalence to the dominance of the more recently-emerged G-associated clades (G, GH, GR) that seem to be accountable for the majority of the cases worldwide since the middle of March 2020. GR, which is the most common isolate clade in Latvia (48.12% of cases), is also a dominant clade in Europe and South America. Currently, GH still seems to be the most common isolate clade circulating throughout North America, but a rise in the number of GR isolates can be observed since the middle of May 2020. The prevalence of GR and, in particular, G clade isolates is also currently on the rise in Africa, and, to a very moderate amount in Oceania and Asia. The relatively high number of isolates not corresponding to any of the currently recognized major SARS-CoV-2 clades (dubbed “Other” or belonging to the “O” clade as of now) in Asia and Oceania makes it possible to speculate about it being indicative of either (but not mutually exclusive), poor quality of the sequences obtained or the possibility of novel clade emergence originating from these regions in the future, should their spread not be effectively contained.

**Figure 2.**
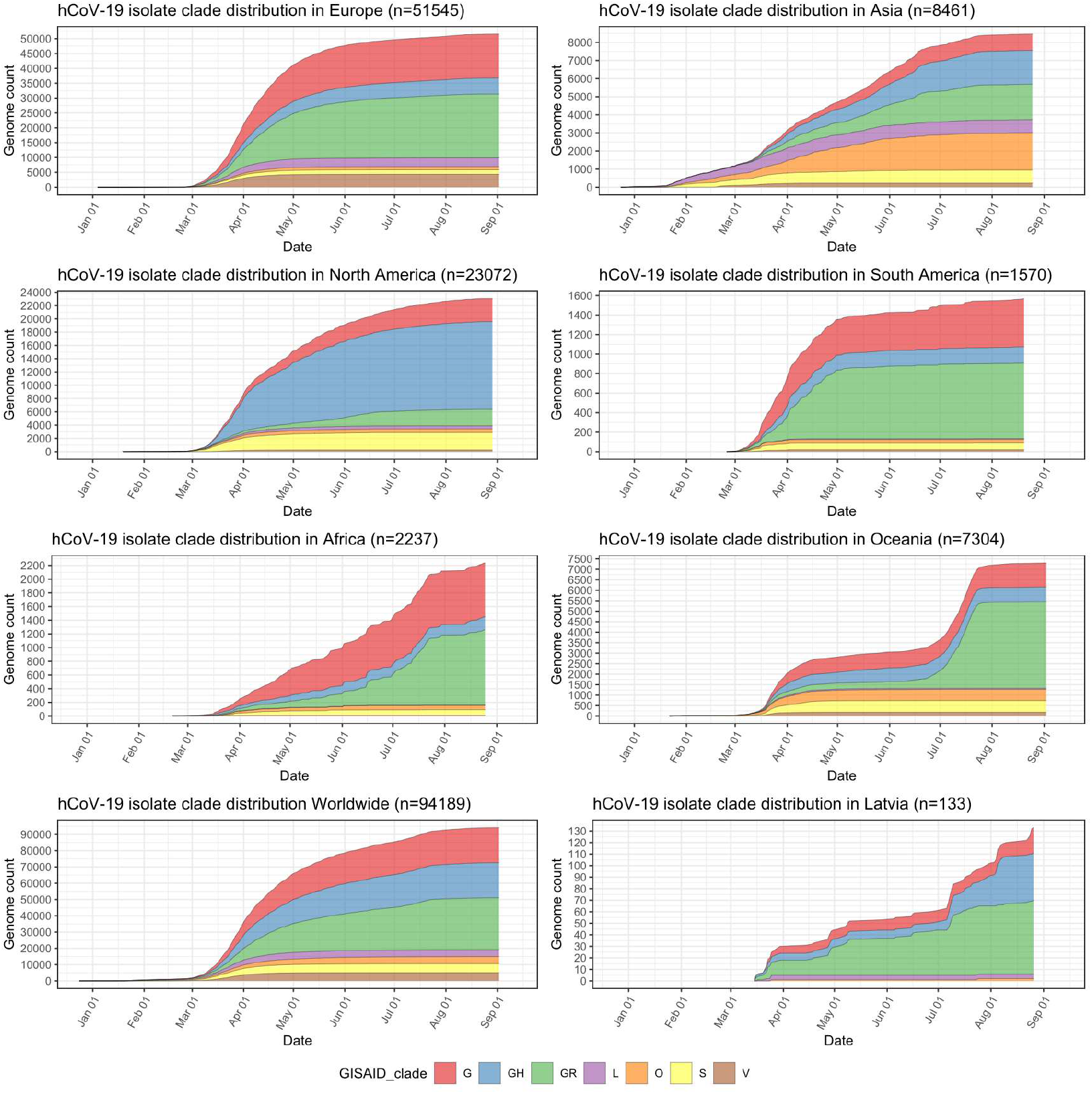
Distribution of sequenced hCoV-19 isolates by clades in major regions of the world, worldwide and in Latvia. Y axis depicts cumulative complete hCoV-19 genome count (with unambiguous collection date) from a particular region and has different scale within the subplots. X axis is the same for all subplots and depicts sampling time-series from 24^th^ of December, 2019 till 12^th^ of September, 2020.

### Mutational landscape of Latvian SARS-CoV-2 isolates

After joining of the neighboring loci, among 133 local isolates, 247 different unique mutational events (154 nonsynonymous, 84 synonymous, 7 substitutions in extragenic regions, single deletion and a single stop codon introduction) that affected 244 positions of the SARS-CoV-2 genome were registered from a total of 1355 variants that were identified. 146 out of 247 distinct mutational events were registered only in one of the 133 samples, while 101 were present in two or more samples (Supplementary table 1 and supplementary figure 2). NSP3 was found to be the mature peptide most frequently affected by non-synonymous substitutions (24 distinct variants resulting in an amino acid change), followed by an N protein that had 15 non-synonymous SNVs documented among Latvian SARS-CoV-2 isolates. Among the most frequently mutated proteins, NSP2, S and NSP12b mature peptides harbored 13, 12 and 10 different amino acid altering mutations, respectively.

Based on the current coronapp web-server [38] report updated at 2020-09-15 (n=89978), most frequent mutational events worldwide are: A23403G corresponding to S:D614G, C3037T silent mutation, C14408T resulting in NSP12b:P314L, C241T extragenic substitution and GGG28881ACC trinucleotide mutation of neighboring loci resulting in N:RG203KR, G25563T – ORF3a:Q57H. All six of these mutations were also among the most frequent mutational events registered in Latvian samples: 5’ UTR C241T extragenic substitution that was present in 129 out of 133 sequenced genomes, while C3037T silent (NSP3:F106F) mutation, A23403G (S:D614G) and C14408T (NSP12b:P314L) were all present in 128/133 samples, GGG28881AAC trinucleotide mutation (N:RG203KR) was observed almost in 59/133 of the samples, and 28881 position of the genome had two more variants detectable in the samples – GGGG28881AACT (N:RG203KL) quadranucleotide mutation (32/133) and G28881A (N:R203K) substitution being present in five of the samples, while G25563T – ORF3a:Q57H was found in 41 of the isolates. (See figure 3 and supplementary table 1).

**Figure 3.**
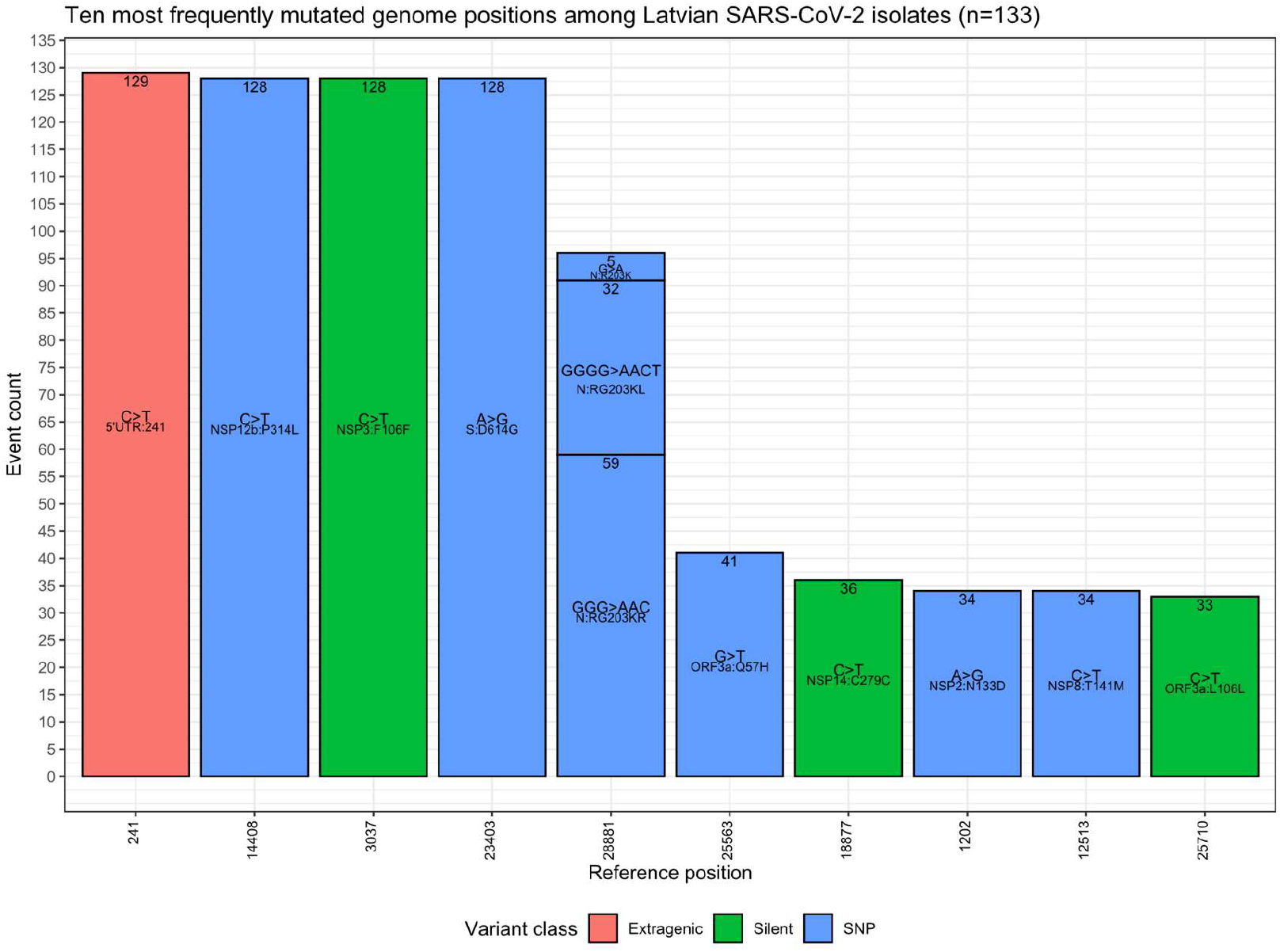
Ten most frequently mutated genome positions among Latvian SARS-CoV-2 isolates (n=133). X axis is discrete and shows genome position corresponding to one of the ten most frequent mutated positions (ordered in descending order). Y axis represents mutation occurrence among the samples (same as numbers within the respective upper boundary of bars) at a given position. Labels in the middle of bars represent the nucleotide change at a given position and its effect on the respective protein amino acid sequence. Color coding is based on the variant class and bars are colored according to the legend. Note: bars representing different mutations of the same locus are stacked (position 28881).

It was noted that five out of six aforementioned mutations (with the exclusion of C14408T) are in the genome positions serving as markers for current SARS-CoV-2 isolate major clade definition and correspond to GR clade, that is the most represented clade Worldwide and hosts almost half of the sequenced isolates in Latvia (Figure 3 and able 1). The C14408T substitution resulting in NSP12b:P314L amino acid change has been previously reported to co-occur with C241T, C3037T and A23403G mutations [51], which is consistent with our data, where four of these SNPs were simultaneously present 127/133 of the Latvian SARS-CoV-2 isolates sequenced up to date. While no experimental evidence of C14408T substitution implications on the NSP12b (RdRp) activity is yet present, isolates bearing this variant were previously speculated to have more mutations, and elaborations about possible implications of RdRp mutations on antiviral drug resistance were made [52]. The fitness of G and G-derived strains, as denoted by the recent rise of their prevalence throughout different regions of the world, is hardly explainable only by the founder effect alone, thus highlighting the fact that further evidence on molecular and clinical implications of the most common substitutions in the genomes of currently circulating SARS-CoV-2 is urgently needed to improve the measures of containment of COVID-19 and develop effective antiviral therapies and vaccines, that would help to not only combat the present virus of immediate concern, but also be of vital importance for other coronaviruses to yet emerge.

### Phylogenetic analyses

Root-to-tip regression analysis with the “best-fitting root” and “correlation” function options resulted in a correlation coefficient of the analysis being estimated at 0.6754 and a determination coefficient (R^2^) equaling to 0.4562 (Supplementary Figure 3). Although having some of the sequences that diverged more or less than expected at their sampling date, the dataset had a moderate association between sequence divergence and sampling date, implying suitability for phylogenetic molecular clock analysis.

Following Bayesian phylogenetic inference, mean mutation rate derived from Latvian SARS-CoV-2 isolates was found to be 7.5185 × 10^−4^ substitutions per site per year (6.0256 × 10^−4^ - 9.1308 × 10^−4^, 95% highest posterior density interval), roughly corresponding to an average of 22-23 mutational events in genome per year (95% HPD: ∼18 – ∼27), and lies within mutation rate ranges predicted by other researchers. Based on the analysis, the estimated most recent common ancestor of the isolates has emerged on 16^th^ of November, 2019 (4^th^ October, 2019 – 27^th^ December, 2019, 95% interval). Our molecular clock analysis (Fig. 4) further supported the more recent divergence of G and G-derived (GR and GH) clades with the most recent common ancestor for three of the aforementioned major clades dating back to 6^th^ of January, 2020 (95% HPD: 27^th^ November, 2019 – 5^th^ February, 2020) and allowed us to date the near-simultaneous emergence of TMRCAs for clades GR (8^th^ of February, 2020; 95% HPD: 16^th^ January, 2020 – 28^th^ of February, 2020) and GH (10^th^ of February, 2020; 95% HPD: 17^th^ January, 2020 – 1^st^ March, 2020). The 95% HPD date ranges are consistent with the collection dates of unambiguously dated genomes belonging to clades GH and GR deposited at GISAID (accessed 2020-08-14). Earliest reported SARS-CoV-2 genome belonging to clade GH was collected on 2nd of February 2020 in Riyadh, Saudi Arabia (GISAID accession: EPI_ISL_489996), while earliest reported GR clade genome was collected on 16^th^ of February 2020 in London, England (GISAID accession: EPI_ISL_466615), however first reported sequences with unambiguous collection date belonging to GR and GH ancestral clade G were collected on 24^th^ of January, 2020 in China, cities of Zhejiang and Chengdu (GISAID accessions: EPI_ISL_422425, EPI_ISL_451345).

**Figure 4.**
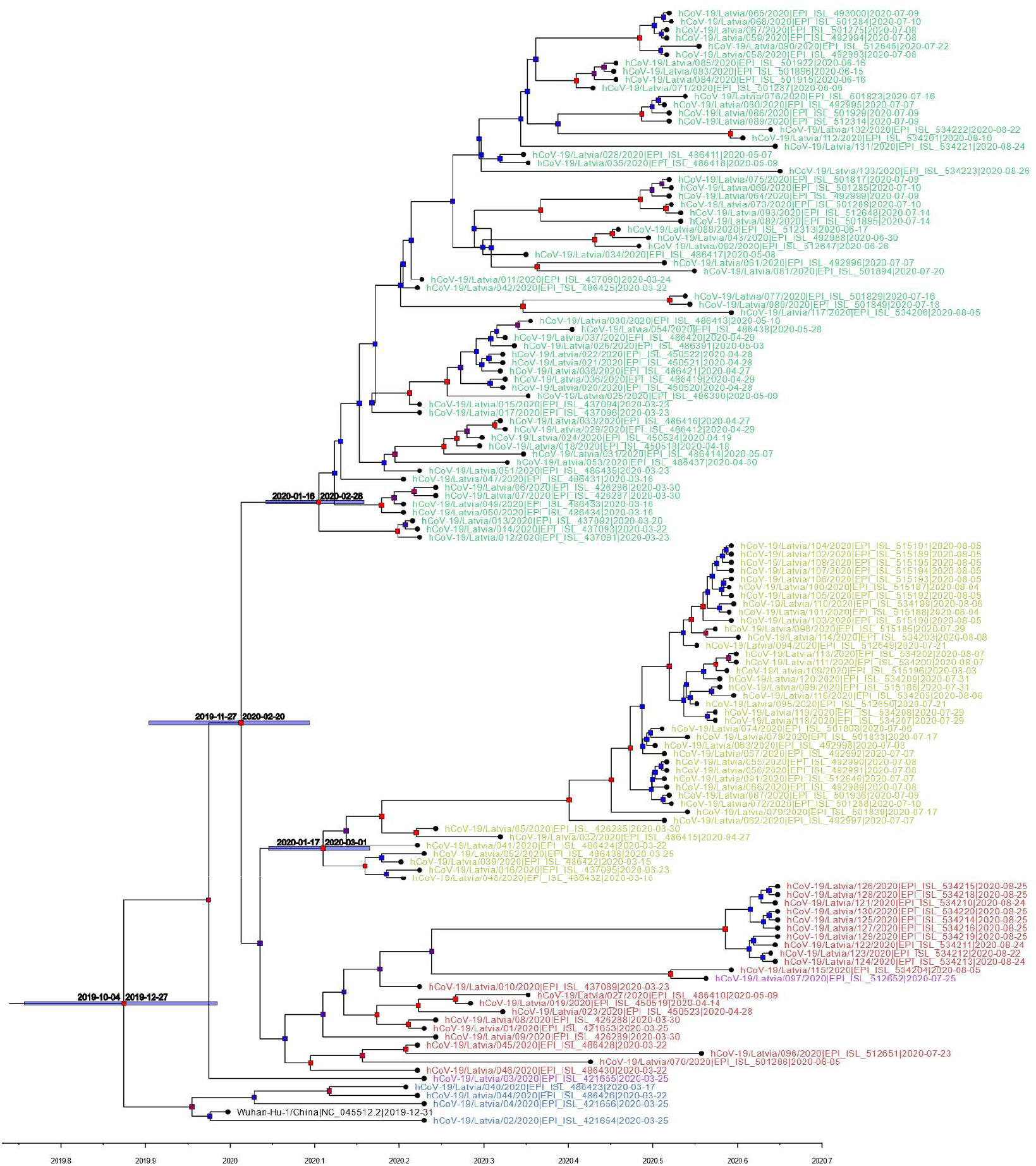
Maximum clade credibility tree (mean node heights) estimated from the completely sequenced Latvian isolates (n=133) and Wuhan-Hu-1 isolate. Node labels are colored according to the GISAID major clade of particular isolate, green – GR, yellow – GH, red – G, blue – L, purple – O (other), black – Wuhan-Hu-1 reference sequence. The tree is timescaled and axis represents time in a decimal year notation (one months is ∼0.08333 of a year and one day is approximately 0.00274 of a year). Nodes are colored according to their respective posterior probabilities in gradient from blue (lowest value) to red (highest value). Dated node bars represent 95% highest posterior density intervals and are shown for the selected nodes.

Our phylogenetic analysis of the local isolates suggests multiple unlinked initial introductions of already divergent SARS-CoV-2 isolates to Latvia. Just two weeks after the first positive case of COVID-19 was documented in Latvia on the 2^nd^ of March, isolates representing at least three major SARS-CoV-2 clades (L, GR and GH) were already circulating within the country corresponding to at least four epidemiologically unlinked introductions. No isolates belonging to clade L (most similar to the initial Wuhan-Hu-1 reference) were sequenced after the end of March and local circulation of clade G representatives was not detectable until the end of August, while clade GH and, specifically, GR isolates seem to have taken hold of the epidemic without showing any signs of ceasing their proliferation within the Latvian population, however, recent reintroduction event possibility should not be ruled out due to cancelation of travel restrictions and insufficient testing of those entering the country. With nearly half of the sequenced isolates belonging to the widely represented GR clade, up to this date, no isolates representing clades V or S were documented among the sequenced Latvian COVID-19 cases (Figures 2 and 4).

Maximum-likelihood phylogenetic tree was built to more apparently infer genetic distances between the samples (Figure 5). Although of satisfactory topology, supporting major clade clustering, the tree evidently shows the possible discrepancies between the reported sampling dates and expected sequence divergence (e.g. some of the samples most divergent from the root are dated with the end of April, while some of the most recently collected ones are notably less divergent), which is not attributable to sequencing errors or the possibility of co-infection by two different “strains”. Identical sequences sampled within a short date range (Figure 5.) might be strongly indicative of epidemiologically linked transmission, given the relatively small daily amount of positive COVID-19 cases in Latvia even during the peaks of the disease spread.

**Figure 5.**
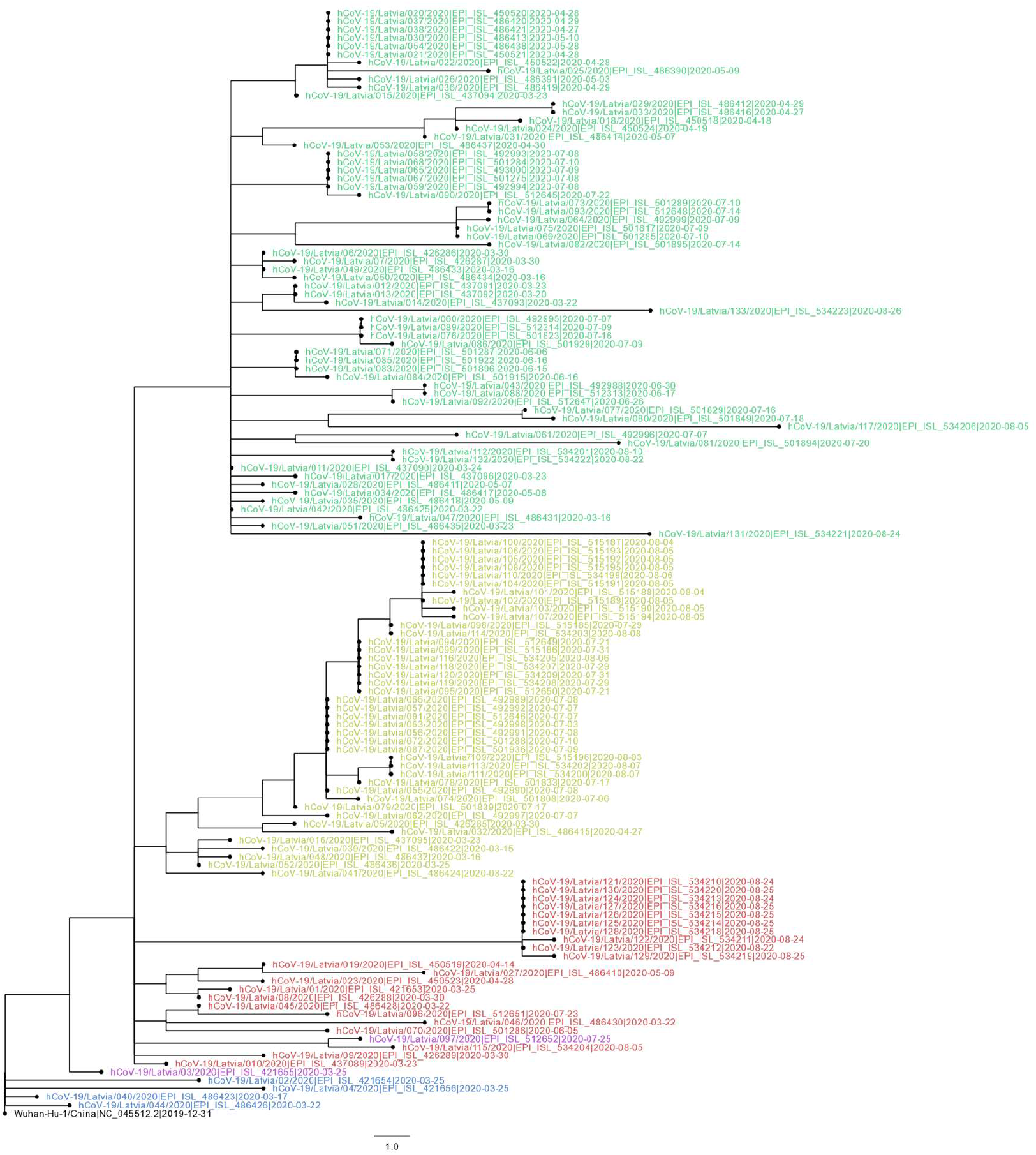
Evolutionary relationships of 133 sequenced Latvian and Wuhan-Hu-1 SARS-CoV-2 isolates. The evolutionary history was inferred using the Maximum-likelihood method allowing for polytomies. The tree is rooted at Wuhan-Hu-1 reference sequence. The tree is drawn to scale, branch lengths correspond to nucleotide substitutions. The analysis involved 134 nucleotide sequences (133 Latvian SARS-CoV-2 isolates and Wuhan-Hu-1 reference sequence). There were a total of 29903 positions in the final dataset. Node labels are colored according to the GISAID major clade of particular isolate, green – GR, yellow – GH, red – G, blue – L, purple – O (other), black – Wuhan-Hu-1 reference sequence.

While providing interesting insights on the COVID-19 situation in Latvia, which might be representative of Baltics region to an extent, given the scarce amount of isolate genomes available from neighboring countries, it, however, should be noted, that the main drawback for each of the presented analyses is stemming from the available dataset - discrete early sampling with some of the dates since first positive case not being sampled at all (Figure 1). Another major drawback is the unavailability of patient/isolate epidemiological data that could be linked to the respective cases sequenced (e.g. sequence epidemiological linkage, patient travel history etc.) in the frame of this study. Inclusion of additional data and retrospective sequencing of a larger number of cases that would allow for a more in-depth analysis of epidemiological situation will be performed and published elsewhere.

In conclusion, the high-throughput framework for SARS-CoV-2 isolate sequencing and data analysis in Latvia has been built by Latvian Biomedical Research and Study Centre early on during the start of the pandemic, tested with the help of both, governmental and local private laboratory sample providers, and proposed as a pivotal tool to monitor the local outbreaks and aid in decision making. This framework allowed us to ensure the sequencing of viral isolates from the majority of the new cases starting from the beginning of July, 2020 with fast date delivery to the Centre for Disease Prevention and Control in Latvia allowing to link the epidemiological data to the isolates being sequenced. We believe that this framework is of vital importance for rapid implementation of the most suitable public health measures, possible transmission history deduction and viral evolution monitoring for the prevention of future epidemiological outbreaks.

## Data Availability

The data used for this study are available either on GISAID or on request to the corresponding author.

https://www.gisaid.org

## Acknowledgements

T he authors acknowledge the use of infrastructure provided by High Performance C omputing Centre of Riga Technical University.

## Ethics statement

The study was approved by the Central Medical Ethics Committee of Latvia (protocol No. 01-29.1/2429 and 01-29.1/1677).

## SUPPLEMENTARY MATERIALS

**Supplementary figure 1.**
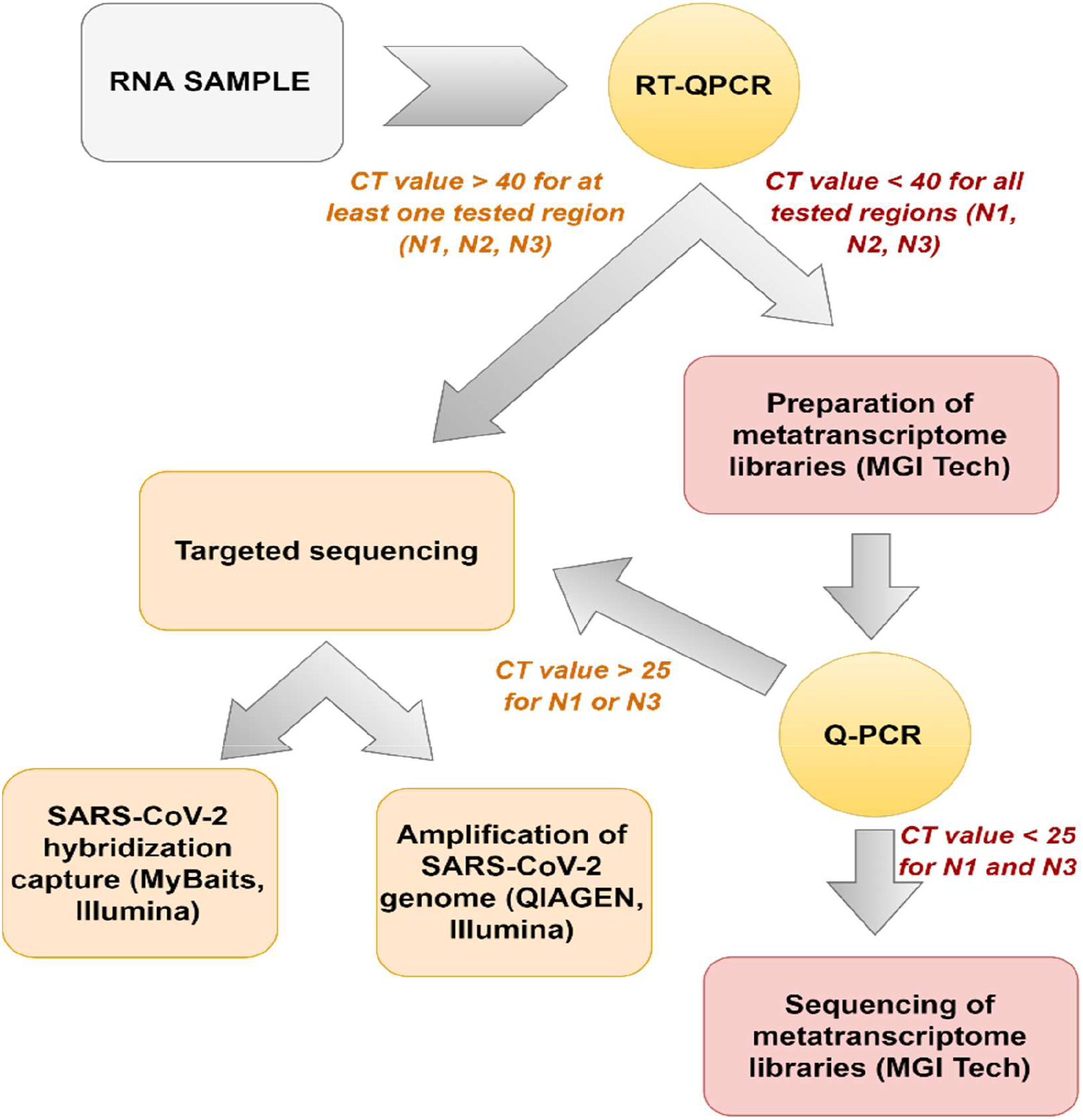
Methodological strategy plan for SARS-CoV-2 genome analysis based on different next-generation sequencing methods.

**Supplementary figure 2.**
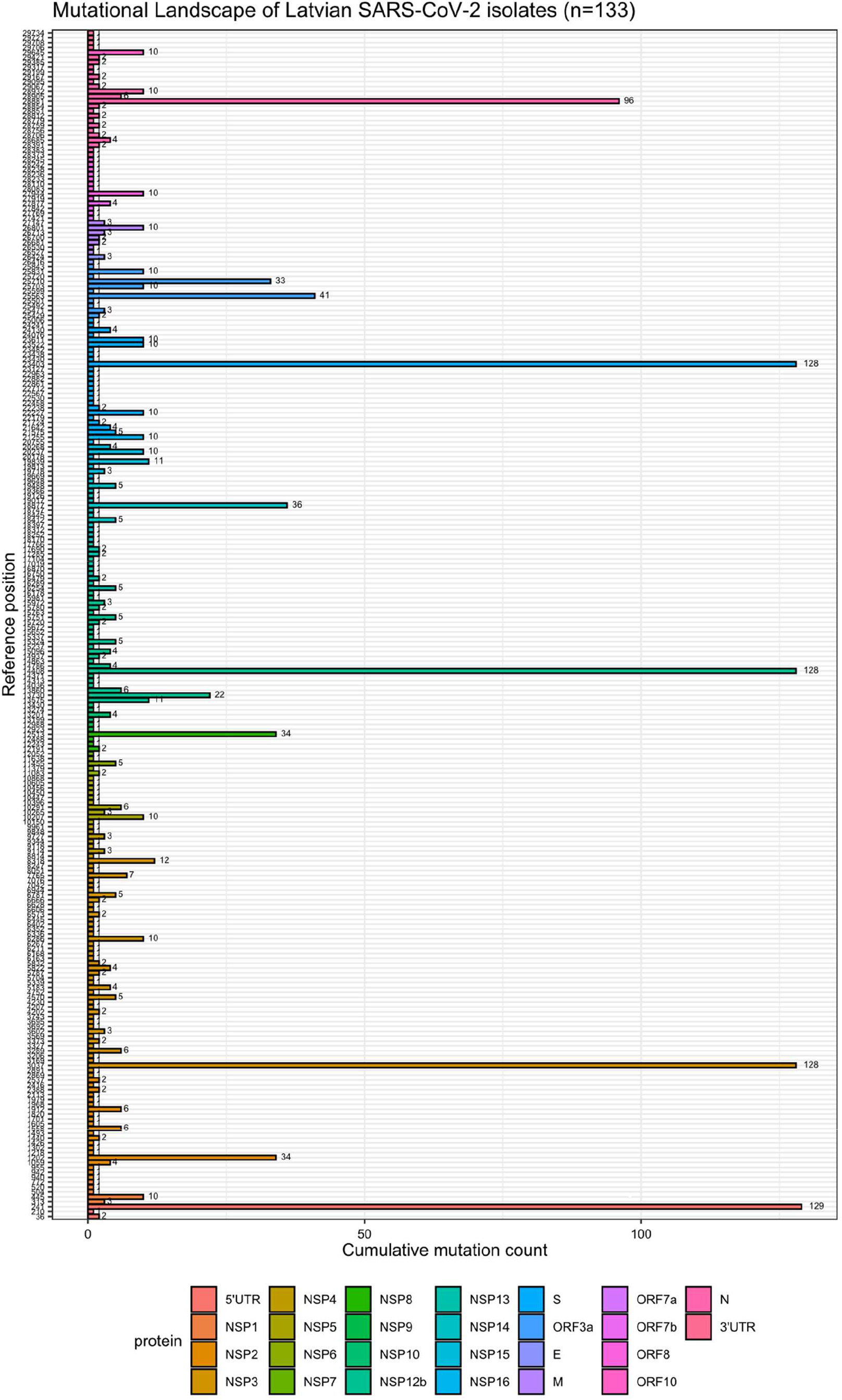
Mutational landscape of Latvian SARS-CoV-2 isolates. Y axis shows the mutated position of a reference SARS-CoV-2 genome. X axis shows the cumulative mutation count at a given position. Number to the right of bars indicate cumulative mutation count at a given position and bars are color-coded according to the protein that the corresponding site participates in encoding. Note, that Y axis is discrete and only positions with mutations documented in local isolates are shown.

**Supplementary figure 3.**
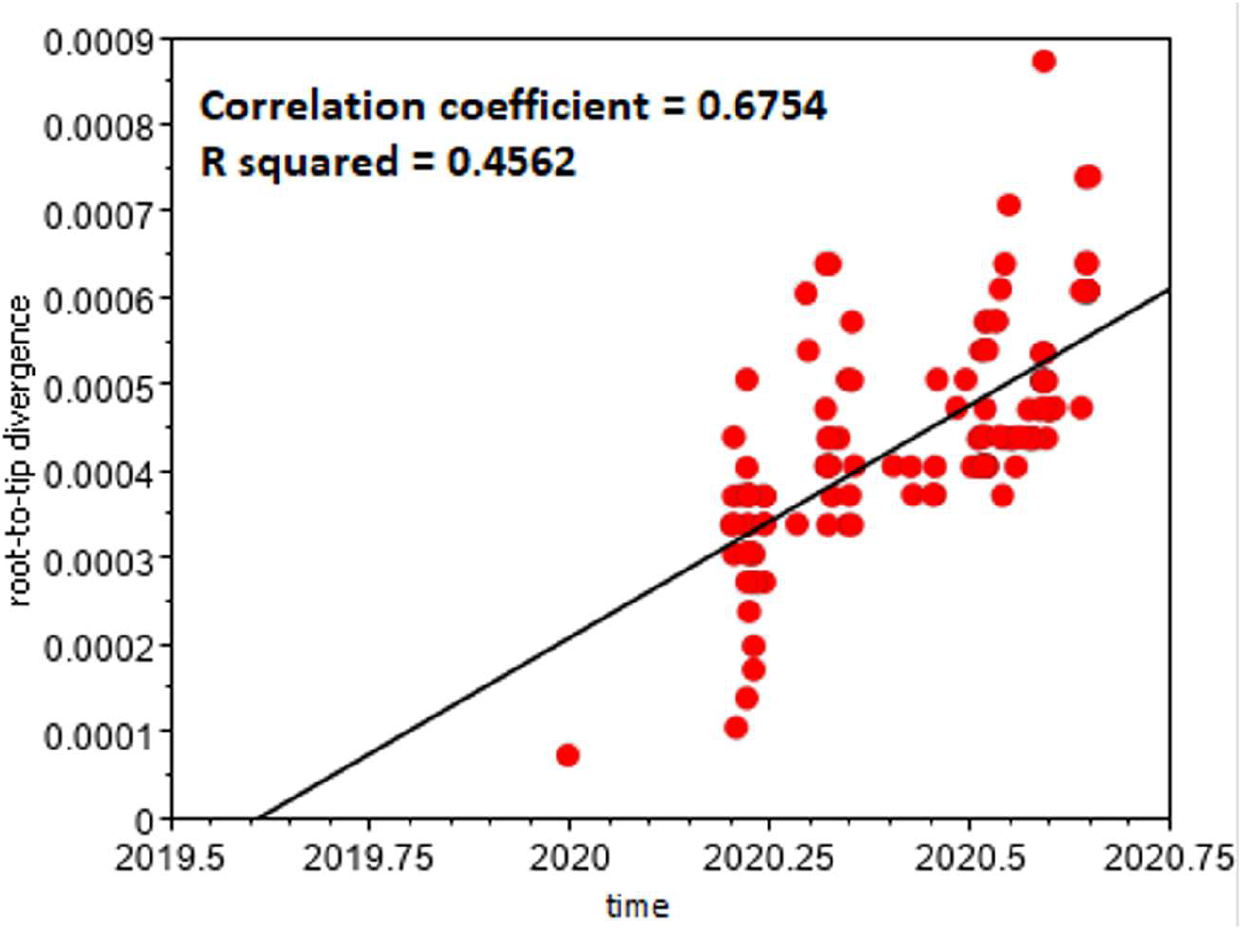
Root-to-tip regression analysis of 133 Latvian SARS-CoV-2 isolates and Wuhan-Hu-1 sequence.

**Supplementary table 1.** Annotation and occurrence of 247 variants identifiable among 133 Latvian SARS-CoV-2 isolates sequenced (sorted by occurrence).

| Variant | Reference | Position in genome | Variant | Variant class | Region affected | Amino acid change | Function of the mature peptide | Variant frequency | Occurrence among Latvian isolates |
| --- | --- | --- | --- | --- | --- | --- | --- | --- | --- |
| 5'UTR:241 | C | 241 | T | Extragenic | 5'UTR | 241 | N/A | 129 | 96.99% |
| NSP3:F106F | C | 3037 | T | Silent | NSP3 | F106F | Predicted phosphoesterase, papain-like proteinase | 128 | 96.24% |
| NSP12b:P314L | C | 14408 | T | SNP | NSP12b | P314L | RNA-dependent RNA polymerase, post-ribosomal frameshift | 128 | 96.24% |
| S:D614G | A | 23403 | G | SNP | S | D614G | Spike | 128 | 96.24% |
| N:RG203KR | GGG | 28881 | AAC | SNP | N | RG203KR | Nucleocapsid protein | 59 | 44.36% |
| ORF3a:Q57H | G | 25563 | T | SNP | ORF3a | Q57H | ORF3a protein | 41 | 30.83% |
| NSP14:C279C | C | 18877 | T | Silent | NSP14 | C279C | 3'-to-5' exonuclease | 36 | 27.07% |
| NSP2:N133D | A | 1202 | G | SNP | NSP2 | N133D | Non-Structural protein 2 | 34 | 25.56% |
| NSP8:T141M | C | 12513 | T | SNP | NSP8 | T141M | Non-Structural Protein 8 | 34 | 25.56% |
| ORF3a:L106L | C | 25710 | T | Silent | ORF3a | L106L | ORF3a protein | 33 | 24.81% |
| N:R203K | G | 28881 | A | SNP | N | R203K | Nucleocapsid protein | 32 | 24.06% |
| NSP12b:A88V | C | 13730 | T | SNP | NSP12b | A88V | RNA-dependent RNA polymerase, post-ribosomal frameshift | 22 | 16.54% |
| NSP3:P1867S | C | 8318 | T | SNP | NSP3 | P1867S | Predicted phosphoesterase, papain-like proteinase | 12 | 9.02% |
| NSP12b:A37S | TG | 13575 | CT | SNP | NSP12b | A37S | RNA-dependent RNA polymerase, post-ribosomal frameshift | 11 | 8.27% |
| NSP15:N73N | T | 19839 | C | Silent | NSP15 | N73N | endoRNase | 11 | 8.27% |
| NSP1:V60V | T | 445 | C | Silent | NSP1 | V60V | Leader protein | 10 | 7.52% |
| NSP3:T1189T | C | 6286 | T | Silent | NSP3 | T1189T | Predicted phosphoesterase, papain-like proteinase | 10 | 7.52% |
| NSP5:N51N | C | 10207 | T | Silent | NSP5 | N51N | 3C-like proteinase | 10 | 7.52% |
| NSP15:R206M | G | 20237 | T | SNP | NSP15 | R206M | endoRNase | 10 | 7.52% |
| NSP16:A199A | G | 21255 | C | Silent | NSP16 | A199A | 2'-O-ribose methyltransferase | 10 | 7.52% |
| S:A222V | C | 22227 | T | SNP | S | A222V | Spike | 10 | 7.52% |
| S:E654K | G | 23522 | A | SNP | S | E654K | Spike | 10 | 7.52% |
| S:R683R | G | 23611 | T | Silent | S | R683R | Spike | 10 | 7.52% |
| ORF3a:P104L | C | 25703 | T | SNP | ORF3a | P104L | ORF3a protein | 10 | 7.52% |
| ORF3a:L147F | C | 25831 | T | SNP | ORF3a | L147F | ORF3a protein | 10 | 7.52% |
| M:L93L | C | 26801 | G | Silent | M | L93L | Membrane | 10 | 7.52% |
| ORF8:H17H | C | 27944 | T | Silent | ORF8 | H17H | ORF8 protein | 10 | 7.52% |
| N:A220V | C | 28932 | T | SNP | N | A220V | Nucleocapsid protein | 10 | 7.52% |
| ORF10:V30L | G | 29645 | T | SNP | ORF10 | V30L | ORF10 protein | 10 | 7.52% |
| NSP3:S1682S | C | 7765 | T | Silent | NSP3 | S1682S | Predicted phosphoesterase, papain-like proteinase | 7 | 5.26% |
| NSP2:I251M | A | 1558 | G | SNP | NSP2 | I251M | Non-Structural protein 2 | 6 | 4.51% |
| NSP3:V190V | T | 3289 | C | Silent | NSP3 | V190V | Predicted phosphoesterase, papain-like proteinase | 6 | 4.51% |
| NSP5:G79G | A | 10291 | G | Silent | NSP5 | G79G | 3C-like proteinase | 6 | 4.51% |
| NSP12b:D131D | C | 13860 | T | Silent | NSP12b | D131D | RNA-dependent RNA polymerase, post-ribosomal frameshift | 6 | 4.51% |
| N:A211V | C | 28905 | T | SNP | N | A211V | Nucleocapsid protein | 6 | 4.51% |
| NSP3:I617I | C | 4570 | T | Silent | NSP3 | I617I | Predicted phosphoesterase, papain-like proteinase | 5 | 3.76% |
| NSP3:F1354F | C | 6781 | T | Silent | NSP3 | F1354F | Predicted phosphoesterase, papain-like proteinase | 5 | 3.76% |
| NSP6:A161A | C | 11455 | T | Silent | NSP6 | A161A | Transmembrane protein | 5 | 3.76% |
| NSP12b:N619N | C | 15324 | T | Silent | NSP12b | N619N | RNA-dependent RNA polymerase, post-ribosomal frameshift | 5 | 3.76% |
| NSP12b:A762S | G | 15751 | T | SNP | NSP12b | A762S | RNA-dependent RNA polymerase, post-ribosomal frameshift | 5 | 3.76% |
| NSP13:V6V | T | 16254 | C | Silent | NSP13 | V6V | Helicase | 5 | 3.76% |
| NSP14:V125F | G | 18412 | T | SNP | NSP14 | V125F | 3'-to-5' exonuclease | 5 | 3.76% |
| NSP14:V483V | C | 19488 | T | Silent | NSP14 | V483V | 3'-to-5' exonuclease | 5 | 3.76% |
| S:L5F | C | 21575 | T | SNP | S | L5F | Spike | 5 | 3.76% |
| N:RG203KL | GGGG | 28881 | AACT | SNP | N | RG203KL | Nucleocapsid protein | 5 | 3.76% |
| NSP2:T85I | C | 1059 | T | SNP | NSP2 | T85I | Non-Structural protein 2 | 4 | 3.01% |
| NSP3:P822S | C | 5183 | T | SNP | NSP3 | P822S | Predicted phosphoesterase, papain-like proteinase | 4 | 3.01% |
| NSP3:L1035F | C | 5822 | T | SNP | NSP3 | L1035F | Predicted phosphoesterase, papain-like proteinase | 4 | 3.01% |
| NSP10:P59P | G | 13201 | A | Silent | NSP10 | P59P | Growth-factor-like protein | 4 | 3.01% |
| NSP12b:A440V | C | 14786 | T | SNP | NSP12b | A440V | RNA-dependent RNA polymerase, post-ribosomal frameshift | 4 | 3.01% |
| NSP12b:N543N | T | 15096 | C | Silent | NSP12b | N543N | RNA-dependent RNA polymerase, post-ribosomal frameshift | 4 | 3.01% |
| NSP15:L216L | A | 20268 | G | Silent | NSP15 | L216L | endoRNase | 4 | 3.01% |
| S:A27V | C | 21642 | T | SNP | S | A27V | Spike | 4 | 3.01% |
| S:N856N | C | 24130 | T | Silent | S | N856N | Spike | 4 | 3.01% |
| ORF7b:C41F | G | 27877 | T | SNP | ORF7b | C41F | ORF7b protein | 4 | 3.01% |
| N:A138S | G | 28685 | T | SNP | N | A138S | Nucleocapsid protein | 4 | 3.01% |
| NSP1:L16L | C | 313 | T | Silent | NSP1 | L16L | Leader protein | 3 | 2.26% |
| NSP3:H295Y | C | 3602 | T | SNP | NSP3 | H295Y | Predicted phosphoesterase, papain-like proteinase | 3 | 2.26% |
| NSP4:P187H | C | 9114 | A | SNP | NSP4 | P187H | Transmembrane protein | 3 | 2.26% |
| NSP4:Y391Y | T | 9727 | C | Silent | NSP4 | Y391Y | Transmembrane protein | 3 | 2.26% |
| NSP5:G71S | G | 10265 | A | SNP | NSP5 | G71S | 3C-like proteinase | 3 | 2.26% |
| NSP12b:V835V | A | 15972 | G | Silent | NSP12b | V835V | RNA-dependent RNA polymerase, post-ribosomal frameshift | 3 | 2.26% |
| NSP15:T33I | C | 19718 | T | SNP | NSP15 | T33I | endoRNase | 3 | 2.26% |
| ORF3a:D27Y | G | 25471 | T | SNP | ORF3a | D27Y | ORF3a protein | 3 | 2.26% |
| E:S60S | T | 26424 | C | Silent | E | S60S | Envelope | 3 | 2.26% |
| M:C64F | G | 26713 | T | SNP | M | C64F | Membrane | 3 | 2.26% |
| M:D209Y | G | 27147 | T | SNP | M | D209Y | Membrane | 3 | 2.26% |
| 5'UTR:36 | C | 36 | G | Extragenic | 5'UTR | 36 | N/A | 2 | 1.50% |
| NSP2:G212D | G | 1440 | A | SNP | NSP2 | G212D | Non-Structural protein 2 | 2 | 1.50% |
| NSP2:S369S | C | 1912 | T | Silent | NSP2 | S369S | Non-Structural protein 2 | 2 | 1.50% |
| NSP2:T528I | C | 2388 | T | SNP | NSP2 | T528I | Non-Structural protein 2 | 2 | 1.50% |
| NSP2:L578V | T | 2537 | G | SNP | NSP2 | L578V | Non-Structural protein 2 | 2 | 1.50% |
| NSP3:D218E | C | 3373 | A | SNP | NSP3 | D218E | Predicted phosphoesterase, papain-like proteinase | 2 | 1.50% |
| NSP3:M494M | C | 4202 | T | Silent | NSP3 | M494M | Predicted phosphoesterase, papain-like proteinase | 2 | 1.50% |
| NSP3:S1023F | C | 5787 | T | SNP | NSP3 | S1023F | Predicted phosphoesterase, papain-like proteinase | 2 | 1.50% |
| NSP3:S1038F | C | 5832 | T | SNP | NSP3 | S1038F | Predicted phosphoesterase, papain-like proteinase | 2 | 1.50% |
| NSP3:S1285F | C | 6573 | T | SNP | NSP3 | S1285F | Predicted phosphoesterase, papain-like proteinase | 2 | 1.50% |
| NSP3:P1316L | C | 6666 | T | SNP | NSP3 | P1316L | Predicted phosphoesterase, papain-like proteinase | 2 | 1.50% |
| NSP6:L37F | G | 11083 | T | SNP | NSP6 | L37F | Transmembrane protein | 2 | 1.50% |
| NSP8:V34F | G | 12191 | T | SNP | NSP8 | V34F | Non-Structural Protein 8 | 2 | 1.50% |
| NSP12b:D490D | C | 14937 | T | Silent | NSP12b | D490D | RNA-dependent RNA polymerase, post-ribosomal frameshift | 2 | 1.50% |
| NSP12b:D751D | C | 15720 | T | Silent | NSP12b | D751D | RNA-dependent RNA polymerase, post-ribosomal frameshift | 2 | 1.50% |
| NSP12b:K771K | G | 15780 | A | Silent | NSP12b | K771K | RNA-dependent RNA polymerase, post-ribosomal frameshift | 2 | 1.50% |
| NSP13:F81F | T | 16479 | C | Silent | NSP13 | F81F | Helicase | 2 | 1.50% |
| NSP13:S350L | C | 17285 | T | SNP | NSP13 | S350L | Helicase | 2 | 1.50% |
| NSP13:S485L | C | 17690 | T | SNP | NSP13 | S485L | Helicase | 2 | 1.50% |
| S:L54L | G | 21724 | A | Silent | S | L54L | Spike | 2 | 1.50% |
| S:L226V | T | 22238 | G | SNP | S | L226V | Spike | 2 | 1.50% |
| ORF3a:V13L | G | 25429 | T | SNP | ORF3a | V13L | ORF3a protein | 2 | 1.50% |
| M:F53F | C | 26681 | T | Silent | M | F53F | Membrane | 2 | 1.50% |
| M:V60L | G | 26700 | T | SNP | M | V60L | Membrane | 2 | 1.50% |
| N:R40C | C | 28391 | T | SNP | N | R40C | Nucleocapsid protein | 2 | 1.50% |
| N:H145Y | C | 28706 | T | SNP | N | H145Y | Nucleocapsid protein | 2 | 1.50% |
| N:P162P | T | 28759 | C | Silent | N | P162P | Nucleocapsid protein | 2 | 1.50% |
| N:S180I | G | 28812 | T | SNP | N | S180I | Nucleocapsid protein | 2 | 1.50% |
| N:S194L | C | 28854 | T | SNP | N | S194L | Nucleocapsid protein | 2 | 1.50% |
| N:T265I | C | 29067 | T | SNP | N | T265I | Nucleocapsid protein | 2 | 1.50% |
| N:Y298Y | C | 29167 | T | Silent | N | Y298Y | Nucleocapsid protein | 2 | 1.50% |
| N:D371G | A | 29385 | G | SNP | N | D371G | Nucleocapsid protein | 2 | 1.50% |
| N:P383L | C | 29421 | T | SNP | N | P383L | Nucleocapsid protein | 2 | 1.50% |
| 5'UTR:210 | G | 210 | T | Extragenic | 5'UTR | 210 | N/A | 1 | 0.75% |
| NSP1:P80L | C | 504 | T | SNP | NSP1 | P80L | Leader protein | 1 | 0.75% |
| NSP1:M85I | G | 520 | T | SNP | NSP1 | M85I | Leader protein | 1 | 0.75% |
| NSP1:V169V | T | 772 | C | Silent | NSP1 | V169V | Leader protein | 1 | 0.75% |
| NSP2:K45K | G | 940 | A | Silent | NSP2 | K45K | Non-Structural protein 2 | 1 | 0.75% |
| NSP2:R46K | G | 942 | A | SNP | NSP2 | R46K | Non-Structural protein 2 | 1 | 0.75% |
| NSP2:C50C | C | 955 | T | Silent | NSP2 | C50C | Non-Structural protein 2 | 1 | 0.75% |
| NSP2:S138L | C | 1218 | T | SNP | NSP2 | S138L | Non-Structural protein 2 | 1 | 0.75% |
| NSP2:T166I | C | 1302 | T | SNP | NSP2 | T166I | Non-Structural protein 2 | 1 | 0.75% |
| NSP2:Y207Y | C | 1426 | T | Silent | NSP2 | Y207Y | Non-Structural protein 2 | 1 | 0.75% |
| NSP2:V230M | G | 1493 | A | SNP | NSP2 | V230M | Non-Structural protein 2 | 1 | 0.75% |
| NSP2:D268 | ATG | 1605 | . | Deletion | NSP2 | D268 | Non-Structural protein 2 | 1 | 0.75% |
| NSP2:S299F | C | 1701 | T | SNP | NSP2 | S299F | Non-Structural protein 2 | 1 | 0.75% |
| NSP2:G339S | G | 1820 | A | SNP | NSP2 | G339S | Non-Structural protein 2 | 1 | 0.75% |
| NSP2:S369S | C | 1912 | A | Silent | NSP2 | S369S | Non-Structural protein 2 | 1 | 0.75% |
| NSP2:T388I | C | 1968 | T | SNP | NSP2 | T388I | Non-Structural protein 2 | 1 | 0.75% |
| NSP2:G392R | G | 1979 | A | SNP | NSP2 | G392R | Non-Structural protein 2 | 1 | 0.75% |
| NSP2:I436I | C | 2113 | T | Silent | NSP2 | I436I | Non-Structural protein 2 | 1 | 0.75% |
| NSP2:Y537Y | C | 2416 | T | Silent | NSP2 | Y537Y | Non-Structural protein 2 | 1 | 0.75% |
| NSP3:V50V | A | 2869 | G | Silent | NSP3 | V50V | Predicted phosphoesterase, papain-like proteinase | 1 | 0.75% |
| NSP3:A58T | G | 2891 | A | SNP | NSP3 | A58T | Predicted phosphoesterase, papain-like proteinase | 1 | 0.75% |
| NSP3:A150A | T | 3169 | C | Silent | NSP3 | A150A | Predicted phosphoesterase, papain-like proteinase | 1 | 0.75% |
| NSP3:D163Y | G | 3206 | T | SNP | NSP3 | D163Y | Predicted phosphoesterase, papain-like proteinase | 1 | 0.75% |
| NSP3:Q203R | A | 3327 | G | SNP | NSP3 | Q203R | Predicted phosphoesterase, papain-like proteinase | 1 | 0.75% |
| NSP3:S284G | A | 3569 | G | SNP | NSP3 | S284G | Predicted phosphoesterase, papain-like proteinase | 1 | 0.75% |
| NSP3:V325F | G | 3692 | T | SNP | NSP3 | V325F | Predicted phosphoesterase, papain-like proteinase | 1 | 0.75% |
| NSP3:V325V | C | 3695 | T | Silent | NSP3 | V325V | Predicted phosphoesterase, papain-like proteinase | 1 | 0.75% |
| NSP3:H342Y | C | 3743 | T | SNP | NSP3 | H342Y | Predicted phosphoesterase, papain-like proteinase | 1 | 0.75% |
| NSP3:A496A | G | 4207 | T | Silent | NSP3 | A496A | Predicted phosphoesterase, papain-like proteinase | 1 | 0.75% |
| NSP3:T504I | C | 4230 | T | SNP | NSP3 | T504I | Predicted phosphoesterase, papain-like proteinase | 1 | 0.75% |
| NSP3:T678I | C | 4752 | T | SNP | NSP3 | T678I | Predicted phosphoesterase, papain-like proteinase | 1 | 0.75% |
| NSP3:P874S | C | 5339 | T | SNP | NSP3 | P874S | Predicted phosphoesterase, papain-like proteinase | 1 | 0.75% |
| NSP3:Q995Q | G | 5704 | A | Silent | NSP3 | Q995Q | Predicted phosphoesterase, papain-like proteinase | 1 | 0.75% |
| NSP3:D1148E | T | 6163 | A | SNP | NSP3 | D1148E | Predicted phosphoesterase, papain-like proteinase | 1 | 0.75% |
| NSP3:V1150A | T | 6168 | C | SNP | NSP3 | V1150A | Predicted phosphoesterase, papain-like proteinase | 1 | 0.75% |
| NSP3:G1164G | A | 6211 | G | Silent | NSP3 | G1164G | Predicted phosphoesterase, papain-like proteinase | 1 | 0.75% |
| NSP3:A1183V | C | 6267 | T | SNP | NSP3 | A1183V | Predicted phosphoesterase, papain-like proteinase | 1 | 0.75% |
| NSP3:S1206L | C | 6336 | T | SNP | NSP3 | S1206L | Predicted phosphoesterase, papain-like proteinase | 1 | 0.75% |
| NSP3:K1211N | G | 6352 | T | SNP | NSP3 | K1211N | Predicted phosphoesterase, papain-like proteinase | 1 | 0.75% |
| NSP3:P1228L | C | 6402 | T | SNP | NSP3 | P1228L | Predicted phosphoesterase, papain-like proteinase | 1 | 0.75% |
| NSP3:D1242D | C | 6445 | T | Silent | NSP3 | D1242D | Predicted phosphoesterase, papain-like proteinase | 1 | 0.75% |
| NSP3:S1296F | C | 6606 | T | SNP | NSP3 | S1296F | Predicted phosphoesterase, papain-like proteinase | 1 | 0.75% |
| NSP3:T1303T | C | 6628 | T | Silent | NSP3 | T1303T | Predicted phosphoesterase, papain-like proteinase | 1 | 0.75% |
| NSP3:I1409V | A | 6944 | G | SNP | NSP3 | I1409V | Predicted phosphoesterase, papain-like proteinase | 1 | 0.75% |
| NSP3:M1441I | G | 7042 | T | SNP | NSP3 | M1441I | Predicted phosphoesterase, papain-like proteinase | 1 | 0.75% |
| NSP3:L1453M | T | 7076 | A | SNP | NSP3 | L1453M | Predicted phosphoesterase, papain-like proteinase | 1 | 0.75% |
| NSP3:N1778D | A | 8051 | G | SNP | NSP3 | N1778D | Predicted phosphoesterase, papain-like proteinase | 1 | 0.75% |
| NSP3:S1843F | C | 8247 | T | SNP | NSP3 | S1843F | Predicted phosphoesterase, papain-like proteinase | 1 | 0.75% |
| NSP4:A87V | C | 8814 | T | SNP | NSP4 | A87V | Transmembrane protein | 1 | 0.75% |
| NSP4:D188D | C | 9118 | T | Silent | NSP4 | D188D | Transmembrane protein | 1 | 0.75% |
| NSP4:L264F | C | 9344 | T | SNP | NSP4 | L264F | Transmembrane protein | 1 | 0.75% |
| NSP4:S432G | A | 9848 | G | SNP | NSP4 | S432G | Transmembrane protein | 1 | 0.75% |
| NSP4:C469C | T | 9961 | C | Silent | NSP4 | C469C | Transmembrane protein | 1 | 0.75% |
| NSP5:L32L | T | 10150 | C | Silent | NSP5 | L32L | 3C-like proteinase | 1 | 0.75% |
| NSP5:V114V | G | 10396 | T | Silent | NSP5 | V114V | 3C-like proteinase | 1 | 0.75% |
| NSP5:R131R | G | 10447 | A | Silent | NSP5 | R131R | 3C-like proteinase | 1 | 0.75% |
| NSP5:P132P | C | 10450 | T | Silent | NSP5 | P132P | 3C-like proteinase | 1 | 0.75% |
| NSP5:F134F | C | 10456 | T | Silent | NSP5 | F134F | 3C-like proteinase | 1 | 0.75% |
| NSP5:P184L | C | 10605 | T | SNP | NSP5 | P184L | 3C-like proteinase | 1 | 0.75% |
| NSP5:L271L | C | 10868 | T | Silent | NSP5 | L271L | 3C-like proteinase | 1 | 0.75% |
| NSP6:A136V | C | 11379 | T | SNP | NSP6 | A136V | Transmembrane protein | 1 | 0.75% |
| NSP6:T222T | T | 11638 | C | Silent | NSP6 | T222T | Transmembrane protein | 1 | 0.75% |
| NSP7:K70N | G | 12052 | T | SNP | NSP7 | K70N | Non-Structural Protein 7 | 1 | 0.75% |
| NSP8:R51L | G | 12243 | T | SNP | NSP8 | R51L | Non-Structural Protein 8 | 1 | 0.75% |
| NSP8:P133S | C | 12488 | T | SNP | NSP8 | P133S | Non-Structural Protein 8 | 1 | 0.75% |
| NSP9:P80T | C | 12923 | A | SNP | NSP9 | P80T | ssRNA-binding protein | 1 | 0.75% |
| NSP9:M101I | G | 12988 | T | SNP | NSP9 | M101I | ssRNA-binding protein | 1 | 0.75% |
| NSP10:P59T | C | 13199 | A | SNP | NSP10 | P59T | Growth-factor-like protein | 1 | 0.75% |
| NSP10:P84S | C | 13274 | T | SNP | NSP10 | P84S | Growth-factor-like protein | 1 | 0.75% |
| NSP10:P136S | C | 13430 | T | SNP | NSP10 | P136S | Growth-factor-like protein | 1 | 0.75% |
| NSP12b:A190D | C | 14036 | A | SNP | NSP12b | A190D | RNA-dependent RNA polymerase, post-ribosomal frameshift | 1 | 0.75% |
| NSP12b:D282D | T | 14313 | C | Silent | NSP12b | D282D | RNA-dependent RNA polymerase, post-ribosomal frameshift | 1 | 0.75% |
| NSP12b:A302S | G | 14371 | T | SNP | NSP12b | A302S | RNA-dependent RNA polymerase, post-ribosomal frameshift | 1 | 0.75% |
| NSP12b:V466I | G | 14863 | A | SNP | NSP12b | V466I | RNA-dependent RNA polymerase, post-ribosomal frameshift | 1 | 0.75% |
| NSP12b:H590H | C | 15237 | T | Silent | NSP12b | H590H | RNA-dependent RNA polymerase, post-ribosomal frameshift | 1 | 0.75% |
| NSP12b:M624V | A | 15337 | G | SNP | NSP12b | M624V | RNA-dependent RNA polymerase, post-ribosomal frameshift | 1 | 0.75% |
| NSP12b:D729Y | G | 15652 | T | SNP | NSP12b | D729Y | RNA-dependent RNA polymerase, post-ribosomal frameshift | 1 | 0.75% |
| NSP12b:E735D | G | 15672 | T | SNP | NSP12b | E735D | RNA-dependent RNA polymerase, post-ribosomal frameshift | 1 | 0.75% |
| NSP12b:G765G | C | 15763 | T | Silent | NSP12b | G765G | RNA-dependent RNA polymerase, post-ribosomal frameshift | 1 | 0.75% |
| NSP12b:I838I | C | 15981 | T | Silent | NSP12b | I838I | RNA-dependent RNA polymerase, post-ribosomal frameshift | 1 | 0.75% |
| NSP12b:S904L | C | 16178 | T | SNP | NSP12b | S904L | RNA-dependent RNA polymerase, post-ribosomal frameshift | 1 | 0.75% |
| NSP13:A18V | C | 16289 | T | SNP | NSP13 | A18V | Helicase | 1 | 0.75% |
| NSP13:P172S | C | 16750 | T | SNP | NSP13 | P172S | Helicase | 1 | 0.75% |
| NSP13:Y211Y | C | 16870 | A | Silent | NSP13 | Y211Y | Helicase | 1 | 0.75% |
| NSP13:E261D | G | 17019 | T | SNP | NSP13 | E261D | Helicase | 1 | 0.75% |
| NSP13:H290Y | C | 17104 | T | SNP | NSP13 | H290Y | Helicase | 1 | 0.75% |
| NSP13:V510V | C | 17766 | T | Silent | NSP13 | V510V | Helicase | 1 | 0.75% |
| NSP14:G44V | G | 18170 | T | SNP | NSP14 | G44V | 3'-to-5' exonuclease | 1 | 0.75% |
| NSP14:N71N | C | 18252 | T | Silent | NSP14 | N71N | 3'-to-5' exonuclease | 1 | 0.75% |
| NSP14:V91V | C | 18312 | T | Silent | NSP14 | V91V | 3'-to-5' exonuclease | 1 | 0.75% |
| NSP14:V120L | G | 18397 | T | SNP | NSP14 | V120L | 3'-to-5' exonuclease | 1 | 0.75% |
| NSP14:V136I | G | 18445 | A | SNP | NSP14 | V136I | 3'-to-5' exonuclease | 1 | 0.75% |
| NSP14:V236V | C | 18747 | T | Silent | NSP14 | V236V | 3'-to-5' exonuclease | 1 | 0.75% |
| NSP14:F326F | C | 19017 | T | Silent | NSP14 | F326F | 3'-to-5' exonuclease | 1 | 0.75% |
| NSP14:I363V | A | 19126 | G | SNP | NSP14 | I363V | 3'-to-5' exonuclease | 1 | 0.75% |
| NSP14:P443S | C | 19366 | T | SNP | NSP14 | P443S | 3'-to-5' exonuclease | 1 | 0.75% |
| NSP15:V10L | G | 19648 | T | SNP | NSP15 | V10L | endoRNase | 1 | 0.75% |
| NSP15:G17R | G | 19669 | A | SNP | NSP15 | G17R | endoRNase | 1 | 0.75% |
| NSP15:P65S | C | 19813 | T | SNP | NSP15 | P65S | endoRNase | 1 | 0.75% |
| NSP15:V186V | C | 20178 | T | Silent | NSP15 | V186V | endoRNase | 1 | 0.75% |
| NSP16:S33R | A | 20755 | C | SNP | NSP16 | S33R | 2'-O-ribose methyltransferase | 1 | 0.75% |
| S:K206R | A | 22179 | G | SNP | S | K206R | Spike | 1 | 0.75% |
| S:T299I | C | 22458 | T | SNP | S | T299I | Spike | 1 | 0.75% |
| S:T323I | C | 22530 | T | SNP | S | T323I | Spike | 1 | 0.75% |
| S:L335L | G | 22567 | A | Silent | S | L335L | Spike | 1 | 0.75% |
| S:P384S | C | 22712 | T | SNP | S | P384S | Spike | 1 | 0.75% |
| S:V433V | T | 22861 | C | Silent | S | V433V | Spike | 1 | 0.75% |
| S:N440K | T | 22882 | G | SNP | S | N440K | Spike | 1 | 0.75% |
| S:D467D | T | 22963 | C | Silent | S | D467D | Spike | 1 | 0.75% |
| S:A522V | C | 23127 | T | SNP | S | A522V | Spike | 1 | 0.75% |
| S:A623V | C | 23430 | T | SNP | S | A623V | Spike | 1 | 0.75% |
| S:A626S | G | 23438 | T | SNP | S | A626S | Spike | 1 | 0.75% |
| S:S640S | T | 23482 | C | Silent | S | S640S | Spike | 1 | 0.75% |
| S:G838G | T | 24076 | C | Silent | S | G838G | Spike | 1 | 0.75% |
| S:A893A | A | 24241 | T | Silent | S | A893A | Spike | 1 | 0.75% |
| S:F1148F | C | 25006 | T | Silent | S | F1148F | Spike | 1 | 0.75% |
| ORF3a:T34A | A | 25492 | G | SNP | ORF3a | T34A | ORF3a protein | 1 | 0.75% |
| ORF3a:I37V | A | 25501 | G | SNP | ORF3a | I37V | ORF3a protein | 1 | 0.75% |
| ORF3a:W69C | G | 25599 | T | SNP | ORF3a | W69C | ORF3a protein | 1 | 0.75% |
| ORF3a:A110S | G | 25720 | T | SNP | ORF3a | A110S | ORF3a protein | 1 | 0.75% |
| ORF3a:T151A | A | 25843 | G | SNP | ORF3a | T151A | ORF3a protein | 1 | 0.75% |
| E:V58F | G | 26416 | T | SNP | E | V58F | Envelope | 1 | 0.75% |
| M:A2V | C | 26527 | T | SNP | M | A2V | Membrane | 1 | 0.75% |
| M:D3G | A | 26530 | G | SNP | M | D3G | Membrane | 1 | 0.75% |
| ORF7a:I10V | A | 27421 | G | SNP | ORF7a | I10V | ORF7a protein | 1 | 0.75% |
| ORF7b:S5L | C | 27769 | T | SNP | ORF7b | S5L | ORF7b protein | 1 | 0.75% |
| ORF7b:W29C | G | 27842 | T | SNP | ORF7b | W29C | ORF7b protein | 1 | 0.75% |
| ORF8:I9T | T | 27919 | C | SNP | ORF8 | I9T | ORF8 protein | 1 | 0.75% |
| ORF8:E64* | G | 28083 | T | Stop introduction | ORF8 | E64* | ORF8 protein | 1 | 0.75% |
| ORF8:Y73H | T | 28110 | C | SNP | ORF8 | Y73H | ORF8 protein | 1 | 0.75% |
| ORF8:V114F | G | 28233 | T | SNP | ORF8 | V114F | ORF8 protein | 1 | 0.75% |
| ORF8:R115C | C | 28236 | T | SNP | ORF8 | R115C | ORF8 protein | 1 | 0.75% |
| ORF8:V116S | TGT | 28238 | ATC | SNP | ORF8 | V116S | ORF8 protein | 1 | 0.75% |
| ORF8:V117L | G | 28242 | C | SNP | ORF8 | V117L | ORF8 protein | 1 | 0.75% |
| ORF8:L118V | T | 28245 | G | SNP | ORF8 | L118V | ORF8 protein | 1 | 0.75% |
| N:G34W | G | 28373 | T | SNP | N | G34W | Nucleocapsid protein | 1 | 0.75% |
| N:S37L | C | 28383 | T | SNP | N | S37L | Nucleocapsid protein | 1 | 0.75% |
| N:L161L | T | 28756 | C | Silent | N | L161L | Nucleocapsid protein | 1 | 0.75% |
| N:K169R | A | 28779 | G | SNP | N | K169R | Nucleocapsid protein | 1 | 0.75% |
| N:S193T | G | 28851 | C | SNP | N | S193T | Nucleocapsid protein | 1 | 0.75% |
| N:F274F | C | 29095 | T | Silent | N | F274F | Nucleocapsid protein | 1 | 0.75% |
| N:P309L | C | 29199 | T | SNP | N | P309L | Nucleocapsid protein | 1 | 0.75% |
| N:D348D | T | 29317 | C | Silent | N | D348D | Nucleocapsid protein | 1 | 0.75% |
| 3'UTR:29706 | G | 29706 | T | Extragenic | 3'UTR | 29706 | N/A | 1 | 0.75% |
| 3'UTR:29708 | C | 29708 | T | Extragenic | 3'UTR | 29708 | N/A | 1 | 0.75% |
| 3'UTR:29721 | C | 29721 | A | Extragenic | 3'UTR | 29721 | N/A | 1 | 0.75% |
| 3'UTR:29734 | G | 29734 | C | Extragenic | 3'UTR | 29734 | N/A | 1 | 0.75% |

**Supplementary table 2.** Latvian SARS-CoV-2 isolate sequencing approaches.

| GISAID isolate name | Accession ID | Collection date | Number of mutations | Average coverage | Type of NGS library | Sequencing technology |
| --- | --- | --- | --- | --- | --- | --- |
| hCoV-19/Latvia/01/2020 | EPI_ISL_421653 | 2020-03-25 | 7 | 7,138x | Metatranscriptome | DNBSEQ-G400RS, MGI Tech Co., Ltd |
| hCoV-19/Latvia/02/2020 | EPI_ISL_421654 | 2020-03-25 | 6 | 91x | Metatranscriptome | DNBSEQ-G400RS, MGI Tech Co., Ltd |
| hCoV-19/Latvia/03/2020 | EPI_ISL_421655 | 2020-03-25 | 3 | 21x | Metatranscriptome | DNBSEQ-G400RS, MGI Tech Co., Ltd |
| hCoV-19/Latvia/04/2020 | EPI_ISL_421656 | 2020-03-25 | 8 | 2,398x | Metatranscriptome | DNBSEQ-G400RS, MGI Tech Co., Ltd |
| hCoV-19/Latvia/05/2020 | EPI_ISL_426285 | 2020-03-30 | 9 | 55x | Metatranscriptome | DNBSEQ-G400RS, MGI Tech Co., Ltd |
| hCoV-19/Latvia/06/2020 | EPI_ISL_426286 | 2020-03-30 | 6 | 1,457x | Metatranscriptome | DNBSEQ-G400RS, MGI Tech Co., Ltd |
| hCoV-19/Latvia/07/2020 | EPI_ISL_426287 | 2020-03-30 | 7 | 129x | Metatranscriptome | DNBSEQ-G400RS, MGI Tech Co., Ltd |
| hCoV-19/Latvia/08/2020 | EPI_ISL_426288 | 2020-03-30 | 6 | 304x | Metatranscriptome | DNBSEQ-G400RS, MGI Tech Co., Ltd |
| hCoV-19/Latvia/09/2020 | EPI_ISL_426289 | 2020-03-30 | 8 | 45x | Metatranscriptome | DNBSEQ-G400RS, MGI Tech Co., Ltd |
| hCoV-19/Latvia/010/2020 | EPI_ISL_437089 | 2020-03-23 | 5 | 518x | Metatranscriptome | DNBSEQ-G400RS, MGI Tech Co., Ltd |
| hCoV-19/Latvia/011/2020 | EPI_ISL_437090 | 2020-03-24 | 5 | 37x | Metatranscriptome | DNBSEQ-G400RS, MGI Tech Co., Ltd |
| hCoV-19/Latvia/012/2020 | EPI_ISL_437091 | 2020-03-23 | 7 | 4,303x | Metatranscriptome | DNBSEQ-G400RS, MGI Tech Co., Ltd |
| hCoV-19/Latvia/013/2020 | EPI_ISL_437092 | 2020-03-20 | 7 | 53x | Metatranscriptome | DNBSEQ-G400RS, MGI Tech Co., Ltd |
| hCoV-19/Latvia/014/2020 | EPI_ISL_437093 | 2020-03-22 | 8 | 511x | Metatranscriptome | DNBSEQ-G400RS, MGI Tech Co., Ltd |
| hCoV-19/Latvia/015/2020 | EPI_ISL_437094 | 2020-03-23 | 6 | 669x | Metatranscriptome | DNBSEQ-G400RS, MGI Tech Co., Ltd |
| hCoV-19/Latvia/016/2020 | EPI_ISL_437095 | 2020-03-23 | 7 | 3,563x | Metatranscriptome | DNBSEQ-G400RS, MGI Tech Co., Ltd |
| hCoV-19/Latvia/017/2020 | EPI_ISL_437096 | 2020-03-23 | 7 | 220x | Metatranscriptome | DNBSEQ-G400RS, MGI Tech Co., Ltd |
| hCoV-19/Latvia/018/2020 | EPI_ISL_450518 | 2020-04-18 | 13 | 2,277x | Metatranscriptome | DNBSEQ-G400RS, MGI Tech Co., Ltd |
| hCoV-19/Latvia/019/2020 | EPI_ISL_450519 | 2020-04-14 | 8 | 6,173x | Metatranscriptome | DNBSEQ-G400RS, MGI Tech Co., Ltd |
| hCoV-19/Latvia/020/2020 | EPI_ISL_450520 | 2020-04-28 | 7 | 1,074x | Metatranscriptome | DNBSEQ-G400RS, MGI Tech Co., Ltd |
| hCoV-19/Latvia/021/2020 | EPI_ISL_450521 | 2020-04-28 | 7 | 563x | Metatranscriptome | DNBSEQ-G400RS, MGI Tech Co., Ltd |
| hCoV-19/Latvia/022/2020 | EPI_ISL_450522 | 2020-04-28 | 8 | 3,836x | Metatranscriptome | DNBSEQ-G400RS, MGI Tech Co., Ltd |
| hCoV-19/Latvia/023/2020 | EPI_ISL_450523 | 2020-04-28 | 8 | 1,363x | Metatranscriptome | DNBSEQ-G400RS, MGI Tech Co., Ltd |
| hCoV-19/Latvia/024/2020 | EPI_ISL_450524 | 2020-04-19 | 11 | 37x | Metatranscriptome | DNBSEQ-G400RS, MGI Tech Co., Ltd |
| hCoV-19/Latvia/025/2020 | EPI_ISL_486390 | 2020-05-09 | 12 | 2715x | Enrichment by hybridization capture | Illumina, MiSeq |
| hCoV-19/Latvia/026/2020 | EPI_ISL_486391 | 2020-05-03 | 8 | 3297x | Enrichment by hybridization capture | Illumina, MiSeq |
| hCoV-19/Latvia/027/2020 | EPI_ISL_486410 | 2020-05-09 | 13 | 5930x | Enrichment by hybridization capture | Illumina, MiSeq |
| hCoV-19/Latvia/028/2020 | EPI_ISL_486411 | 2020-05-07 | 6 | 191x | Metatranscriptome | DNBSEQ-G400RS, MGI Tech Co., Ltd |
| hCoV-19/Latvia/029/2020 | EPI_ISL_486412 | 2020-04-29 | 14 | 7839x | Metatranscriptome | DNBSEQ-G400RS, MGI Tech Co., Ltd |
| hCoV-19/Latvia/030/2020 | EPI_ISL_486413 | 2020-05-10 | 7 | 861x | Metatranscriptome | DNBSEQ-G400RS, MGI Tech Co., Ltd |
| hCoV-19/Latvia/031/2020 | EPI_ISL_486414 | 2020-05-07 | 10 | 79x | Metatranscriptome | DNBSEQ-G400RS, MGI Tech Co., Ltd |
| hCoV-19/Latvia/032/2020 | EPI_ISL_486415 | 2020-04-27 | 12 | 330x | Metatranscriptome | DNBSEQ-G400RS, MGI Tech Co., Ltd |
| hCoV-19/Latvia/033/2020 | EPI_ISL_486416 | 2020-04-27 | 14 | 27x | Metatranscriptome | DNBSEQ-G400RS, MGI Tech Co., Ltd |
| hCoV-19/Latvia/034/2020 | EPI_ISL_486417 | 2020-05-08 | 7 | 138x | Metatranscriptome | DNBSEQ-G400RS, MGI Tech Co., Ltd |
| hCoV-19/Latvia/035/2020 | EPI_ISL_486418 | 2020-05-09 | 6 | 604x | Metatranscriptome | DNBSEQ-G400RS, MGI Tech Co., Ltd |
| hCoV-19/Latvia/036/2020 | EPI_ISL_486419 | 2020-04-29 | 8 | 25x | Metatranscriptome | DNBSEQ-G400RS, MGI Tech Co., Ltd |
| hCoV-19/Latvia/037/2020 | EPI_ISL_486420 | 2020-04-29 | 7 | 30x | Metatranscriptome | DNBSEQ-G400RS, MGI Tech Co., Ltd |
| hCoV-19/Latvia/038/2020 | EPI_ISL_486421 | 2020-04-27 | 7 | 7480x | Metatranscriptome | DNBSEQ-G400RS, MGI Tech Co., Ltd |
| hCoV-19/Latvia/039/2020 | EPI_ISL_486422 | 2020-03-15 | 8 | 6066x | Enrichment by hybridization capture | Illumina, MiSeq |
| hCoV-19/Latvia/040/2020 | EPI_ISL_486423 | 2020-03-17 | 1 | 7129x | Enrichment by hybridization capture | Illumina, MiSeq |
| hCoV-19/Latvia/041/2020 | EPI_ISL_486424 | 2020-03-22 | 8 | 6560x | Enrichment by hybridization capture | Illumina, MiSeq |
| hCoV-19/Latvia/042/2020 | EPI_ISL_486425 | 2020-03-22 | 5 | 4274x | Enrichment by hybridization capture | Illumina, MiSeq |
| hCoV-19/Latvia/044/2020 | EPI_ISL_486426 | 2020-06-30 | 11 | 2763x | Enrichment by hybridization capture | Illumina, MiSeq |
| hCoV-19/Latvia/045/2020 | EPI_ISL_486428 | 2020-03-22 | 2 | 4545x | Enrichment by hybridization capture | Illumina, MiSeq |
| hCoV-19/Latvia/046/2020 | EPI_ISL_486430 | 2020-03-22 | 6 | 4368x | Enrichment by hybridization capture | Illumina, MiSeq |
| hCoV-19/Latvia/047/2020 | EPI_ISL_486431 | 2020-03-22 | 13 | 6833x | Enrichment by hybridization capture | Illumina, MiSeq |
| hCoV-19/Latvia/048/2020 | EPI_ISL_486432 | 2020-03-16 | 9 | 6247x | Enrichment by hybridization capture | Illumina, MiSeq |
| hCoV-19/Latvia/049/2020 | EPI_ISL_486433 | 2020-03-16 | 7 | 5337x | Enrichment by hybridization capture | Illumina, MiSeq |
| hCoV-19/Latvia/050/2020 | EPI_ISL_486434 | 2020-03-16 | 6 | 6222x | Enrichment by hybridization capture | Illumina, MiSeq |
| hCoV-19/Latvia/051/2020 | EPI_ISL_486435 | 2020-03-16 | 7 | 4398x | Enrichment by hybridization capture | Illumina, MiSeq |
| hCoV-19/Latvia/052/2020 | EPI_ISL_486436 | 2020-03-23 | 6 | 846x | Enrichment by hybridization capture | Illumina, MiSeq |
| hCoV-19/Latvia/053/2020 | EPI_ISL_486437 | 2020-03-25 | 6 | 43x | Metatranscriptome | DNBSEQ-G400RS, MGI Tech Co., Ltd |
| hCoV-19/Latvia/054/2020 | EPI_ISL_486438 | 2020-04-30 | 7 | 63x | Metatranscriptome | DNBSEQ-G400RS, MGI Tech Co., Ltd |
| hCoV-19/Latvia/043/2020 | EPI_ISL_492988 | 2020-05-28 | 7 | 3991x | Amplification by multiplexed primers | Illumina, MiSeq |
| hCoV-19/Latvia/066/2020 | EPI_ISL_492989 | 2020-07-08 | 10 | 4404x | Amplification by multiplexed primers | Illumina, MiSeq |
| hCoV-19/Latvia/055/2020 | EPI_ISL_492990 | 2020-07-08 | 10 | 3715x | Amplification by multiplexed primers | Illumina, MiSeq |
| hCoV-19/Latvia/056/2020 | EPI_ISL_492991 | 2020-07-07 | 10 | 3801x | Amplification by multiplexed primers | Illumina, MiSeq |
| hCoV-19/Latvia/057/2020 | EPI_ISL_492992 | 2020-07-08 | 8 | 4169x | Amplification by multiplexed primers | Illumina, MiSeq |
| hCoV-19/Latvia/058/2020 | EPI_ISL_492993 | 2020-07-08 | 8 | 4322x | Amplification by multiplexed primers | Illumina, MiSeq |
| hCoV-19/Latvia/059/2020 | EPI_ISL_492994 | 2020-07-07 | 9 | 3363x | Amplification by multiplexed primers | Illumina, MiSeq |
| hCoV-19/Latvia/060/2020 | EPI_ISL_492995 | 2020-07-07 | 12 | 4163x | Amplification by multiplexed primers | Illumina, MiSeq |
| hCoV-19/Latvia/061/2020 | EPI_ISL_492996 | 2020-07-07 | 10 | 3534x | Amplification by multiplexed primers | Illumina, MiSeq |
| hCoV-19/Latvia/062/2020 | EPI_ISL_492997 | 2020-07-03 | 10 | 1334x | Amplification by multiplexed primers | Illumina, MiSeq |
| hCoV-19/Latvia/063/2020 | EPI_ISL_492998 | 2020-07-09 | 13 | 4315x | Amplification by multiplexed primers | Illumina, MiSeq |
| hCoV-19/Latvia/064/2020 | EPI_ISL_492999 | 2020-07-09 | 8 | 773x | Amplification by multiplexed primers | Illumina, MiSeq |
| hCoV-19/Latvia/065/2020 | EPI_ISL_493000 | 2020-07-08 | 10 | 4022x | Amplification by multiplexed primers | Illumina, MiSeq |
| hCoV-19/Latvia/067/2020 | EPI_ISL_501275 | 2020-07-08 | 8 | 3200x | Amplification by multiplexed primers | Illumina, MiSeq |
| hCoV-19/Latvia/068/2020 | EPI_ISL_501284 | 2020-07-10 | 8 | 3682x | Amplification by multiplexed primers | Illumina, MiSeq |
| hCoV-19/Latvia/069/2020 | EPI_ISL_501285 | 2020-07-10 | 12 | 4184x | Amplification by multiplexed primers | Illumina, MiSeq |
| hCoV-19/Latvia/070/2020 | EPI_ISL_501286 | 2020-06-05 | 10 | 3932x | Amplification by multiplexed primers | Illumina, MiSeq |
| hCoV-19/Latvia/071/2020 | EPI_ISL_501287 | 2020-06-06 | 7 | 3345x | Amplification by multiplexed primers | Illumina, MiSeq |
| hCoV-19/Latvia/072/2020 | EPI_ISL_501288 | 2020-07-10 | 10 | 4639x | Amplification by multiplexed primers | Illumina, MiSeq |
| hCoV-19/Latvia/073/2020 | EPI_ISL_501289 | 2020-07-10 | 13 | 4606x | Amplification by multiplexed primers | Illumina, MiSeq |
| hCoV-19/Latvia/074/2020 | EPI_ISL_501808 | 2020-07-06 | 11 | 5190x | Amplification by multiplexed primers | Illumina, MiSeq |
| hCoV-19/Latvia/075/2020 | EPI_ISL_501817 | 2020-07-09 | 12 | 2923x | Amplification by multiplexed primers | Illumina, MiSeq |
| hCoV-19/Latvia/076/2020 | EPI_ISL_501823 | 2020-07-16 | 9 | 3816x | Amplification by multiplexed primers | Illumina, MiSeq |
| hCoV-19/Latvia/077/2020 | EPI_ISL_501829 | 2020-07-16 | 14 | 1132x | Amplification by multiplexed primers | Illumina, MiSeq |
| hCoV-19/Latvia/078/2020 | EPI_ISL_501833 | 2020-07-17 | 11 | 4088x | Amplification by multiplexed primers | Illumina, MiSeq |
| hCoV-19/Latvia/079/2020 | EPI_ISL_501839 | 2020-07-17 | 9 | 322x | Amplification by multiplexed primers | Illumina, MiSeq |
| hCoV-19/Latvia/080/2020 | EPI_ISL_501849 | 2020-07-18 | 15 | 2729x | Amplification by multiplexed primers | Illumina, MiSeq |
| hCoV-19/Latvia/081/2020 | EPI_ISL_501894 | 2020-07-20 | 15 | 250x | Amplification by multiplexed primers | Illumina, MiSeq |
| hCoV-19/Latvia/082/2020 | EPI_ISL_501895 | 2020-07-14 | 13 | 4873x | Amplification by multiplexed primers | Illumina, MiSeq |
| hCoV-19/Latvia/083/2020 | EPI_ISL_501896 | 2020-06-15 | 7 | 5512x | Amplification by multiplexed primers | Illumina, MiSeq |
| hCoV-19/Latvia/084/2020 | EPI_ISL_501915 | 2020-06-16 | 8 | 4766x | Amplification by multiplexed primers | Illumina, MiSeq |
| hCoV-19/Latvia/085/2020 | EPI_ISL_501922 | 2020-06-16 | 7 | 3296x | Amplification by multiplexed primers | Illumina, MiSeq |
| hCoV-19/Latvia/086/2020 | EPI_ISL_501929 | 2020-07-09 | 10 | 4980x | Amplification by multiplexed primers | Illumina, MiSeq |
| hCoV-19/Latvia/087/2020 | EPI_ISL_501936 | 2020-07-09 | 10 | 4248x | Amplification by multiplexed primers | Illumina, MiSeq |
| hCoV-19/Latvia/088/2020 | EPI_ISL_512313 | 2020-06-17 | 11 | 2883x | Amplification by multiplexed primers | Illumina, MiSeq |
| hCoV-19/Latvia/089/2020 | EPI_ISL_512314 | 2020-07-09 | 9 | 3332x | Amplification by multiplexed primers | Illumina, MiSeq |
| hCoV-19/Latvia/090/2020 | EPI_ISL_512645 | 2020-07-22 | 9 | 3330x | Amplification by multiplexed primers | Illumina, MiSeq |
| hCoV-19/Latvia/091/2020 | EPI_ISL_512646 | 2020-07-07 | 10 | 3593x | Amplification by multiplexed primers | Illumina, MiSeq |
| hCoV-19/Latvia/092/2020 | EPI_ISL_512647 | 2020-06-26 | 10 | 2643x | Amplification by multiplexed primers | Illumina, MiSeq |
| hCoV-19/Latvia/093/2020 | EPI_ISL_512648 | 2020-07-14 | 13 | 3664x | Amplification by multiplexed primers | Illumina, MiSeq |
| hCoV-19/Latvia/094/2020 | EPI_ISL_512649 | 2020-07-21 | 11 | 3682x | Amplification by multiplexed primers | Illumina, MiSeq |
| hCoV-19/Latvia/095/2020 | EPI_ISL_512650 | 2020-07-21 | 11 | 3745x | Amplification by multiplexed primers | Illumina, MiSeq |
| hCoV-19/Latvia/096/2020 | EPI_ISL_512651 | 2020-07-23 | 10 | 3320x | Amplification by multiplexed primers | Illumina, MiSeq |
| hCoV-19/Latvia/097/2020 | EPI_ISL_512652 | 2020-07-25 | 9 | 2830x | Amplification by multiplexed primers | Illumina, MiSeq |
| hCoV-19/Latvia/098/2020 | EPI_ISL_515185 | 2020-07-29 | 12 | 4176x | Amplification by multiplexed primers | Illumina, MiSeq |
| hCoV-19/Latvia/099/2020 | EPI_ISL_515186 | 2020-07-31 | 11 | 3191x | Amplification by multiplexed primers | Illumina, MiSeq |
| hCoV-19/Latvia/100/2020 | EPI_ISL_515187 | 2020-08-04 | 13 | 4296x | Amplification by multiplexed primers | Illumina, MiSeq |
| hCoV-19/Latvia/101/2020 | EPI_ISL_515188 | 2020-08-04 | 14 | 4137x | Amplification by multiplexed primers | Illumina, MiSeq |
| hCoV-19/Latvia/102/2020 | EPI_ISL_515189 | 2020-08-05 | 13 | 3888x | Amplification by multiplexed primers | Illumina, MiSeq |
| hCoV-19/Latvia/103/2020 | EPI_ISL_515190 | 2020-08-05 | 14 | 3488x | Amplification by multiplexed primers | Illumina, MiSeq |
| hCoV-19/Latvia/104/2020 | EPI_ISL_515191 | 2020-08-05 | 13 | 4111x | Amplification by multiplexed primers | Illumina, MiSeq |
| hCoV-19/Latvia/105/2020 | EPI_ISL_515192 | 2020-08-05 | 13 | 3429x | Amplification by multiplexed primers | Illumina, MiSeq |
| hCoV-19/Latvia/106/2020 | EPI_ISL_515193 | 2020-08-05 | 13 | 4349x | Amplification by multiplexed primers | Illumina, MiSeq |
| hCoV-19/Latvia/107/2020 | EPI_ISL_515194 | 2020-08-05 | 14 | 3860x | Amplification by multiplexed primers | Illumina, MiSeq |
| hCoV-19/Latvia/108/2020 | EPI_ISL_515195 | 2020-08-05 | 13 | 3696x | Amplification by multiplexed primers | Illumina, MiSeq |
| hCoV-19/Latvia/109/2020 | EPI_ISL_515196 | 2020-08-03 | 12 | 1090x | Amplification by multiplexed primers | Illumina, MiSeq |
| hCoV-19/Latvia/110/2020 | EPI_ISL_534199 | 2020-08-06 | 13 | 3711x | Amplification by multiplexed primers | Illumina, MiSeq |
| hCoV-19/Latvia/111/2020 | EPI_ISL_534200 | 2020-08-07 | 12 | 4159x | Amplification by multiplexed primers | Illumina, MiSeq |
| hCoV-19/Latvia/112/2020 | EPI_ISL_534201 | 2020-08-10 | 10 | 2691x | Amplification by multiplexed primers | Illumina, MiSeq |
| hCoV-19/Latvia/113/2020 | EPI_ISL_534202 | 2020-08-07 | 12 | 3601x | Amplification by multiplexed primers | Illumina, MiSeq |
| hCoV-19/Latvia/114/2020 | EPI_ISL_534203 | 2020-08-08 | 12 | 1822x | Amplification by multiplexed primers | Illumina, MiSeq |
| hCoV-19/Latvia/115/2020 | EPI_ISL_534204 | 2020-08-05 | 12 | 2597x | Amplification by multiplexed primers | Illumina, MiSeq |
| hCoV-19/Latvia/116/2020 | EPI_ISL_534205 | 2020-08-06 | 11 | 3587x | Amplification by multiplexed primers | Illumina, MiSeq |
| hCoV-19/Latvia/117/2020 | EPI_ISL_534206 | 2020-08-05 | 22 | 3284x | Amplification by multiplexed primers | Illumina, MiSeq |
| hCoV-19/Latvia/118/2020 | EPI_ISL_534207 | 2020-07-29 | 11 | 3251x | Amplification by multiplexed primers | Illumina, MiSeq |
| hCoV-19/Latvia/119/2020 | EPI_ISL_534208 | 2020-07-29 | 11 | 2955x | Amplification by multiplexed primers | Illumina, MiSeq |
| hCoV-19/Latvia/120/2020 | EPI_ISL_534209 | 2020-07-31 | 11 | 2050x | Amplification by multiplexed primers | Illumina, MiSeq |
| hCoV-19/Latvia/121/2020 | EPI_ISL_534210 | 2020-08-24 | 16 | 2283x | Amplification by multiplexed primers | Illumina, MiSeq |
| hCoV-19/Latvia/122/2020 | EPI_ISL_534211 | 2020-08-24 | 17 | 3480x | Amplification by multiplexed primers | Illumina, MiSeq |
| hCoV-19/Latvia/123/2020 | EPI_ISL_534212 | 2020-08-22 | 16 | 321x | Amplification by multiplexed primers | Illumina, MiSeq |
| hCoV-19/Latvia/124/2020 | EPI_ISL_534213 | 2020-08-24 | 16 | 643x | Amplification by multiplexed primers | Illumina, MiSeq |
| hCoV-19/Latvia/125/2020 | EPI_ISL_534214 | 2020-08-25 | 16 | 3986x | Amplification by multiplexed primers | Illumina, MiSeq |
| hCoV-19/Latvia/126/2020 | EPI_ISL_534215 | 2020-08-25 | 16 | 2327x | Amplification by multiplexed primers | Illumina, MiSeq |
| hCoV-19/Latvia/127/2020 | EPI_ISL_534216 | 2020-08-25 | 16 | 3292x | Amplification by multiplexed primers | Illumina, MiSeq |
| hCoV-19/Latvia/128/2020 | EPI_ISL_534218 | 2020-08-25 | 16 | 3387x | Amplification by multiplexed primers | Illumina, MiSeq |
| hCoV-19/Latvia/129/2020 | EPI_ISL_534219 | 2020-08-25 | 17 | 3125x | Amplification by multiplexed primers | Illumina, MiSeq |
| hCoV-19/Latvia/130/2020 | EPI_ISL_534220 | 2020-08-25 | 16 | 3804x | Amplification by multiplexed primers | Illumina, MiSeq |
| hCoV-19/Latvia/131/2020 | EPI_ISL_534221 | 2020-08-24 | 18 | 3702x | Amplification by multiplexed primers | Illumina, MiSeq |
| hCoV-19/Latvia/132/2020 | EPI_ISL_534222 | 2020-08-22 | 10 | 3324x | Amplification by multiplexed primers | Illumina, MiSeq |
| hCoV-19/Latvia/133/2020 | EPI_ISL_534223 | 2020-08-26 | 18 | 3588x | Amplification by multiplexed primers | Illumina, MiSeq |

